# Efficacy of COVID-19 vaccines in immunocompromised patients: A systematic review and meta-analysis

**DOI:** 10.1101/2021.09.28.21264126

**Authors:** Ainsley Ryan Yan Bin Lee, Shi Yin Wong, Louis Yi Ann Chai, Soo Chin Lee, Matilda Lee, Mark Dhinesh Muthiah, Sen Hee Tay, Chong Boon Teo, Benjamin Kye Jyn Tan, Yiong Huak Chan, Raghav Sundar, Yu Yang Soon

## Abstract

**Objective:** To compare the efficacy of COVID 19 vaccines between those with immunocompromised medical conditions and those who are immunocompetent

**Design:** Systematic review and meta-analysis

**Data sources:** PubMed, EMBASE, CENTRAL, CORD-19 and WHO COVID-19 research databases were searched for eligible comparative studies published between 1 December 2020 and 3 September 2021. ClinicalTrials.gov and the WHO International Clinical Trials Registry Platform were searched in September 2021 to identify registered yet unpublished or ongoing studies.

**Study selection:** Prospective observational studies which compared the efficacy of COVID-19 vaccination between those with immunocompromising medical conditions and those who were immunocompetent were included. Two reviewers independently screened for potentially eligible studies.

**Data extraction:** The primary outcomes of interest were cumulative incidence of seroconversion after first and second doses of COVID vaccination. Secondary outcomes included SARS-CoV-2 antibody titre level after first and second doses of COVID-19 vaccination. After duplicate data abstraction, a frequentist random effects meta-analysis was conducted. Risk of bias was assessed using the ROBINS-I tool. Certainty of evidence was assessed using the GRADE approach.

**Results:** After screening 3283 studies, 42 studies that met our inclusion criteria were identified. 18 immunocompromised cohorts from 17 studies reported seroconversion in immunocompromised patients compared to healthy controls after the first dose and 30 immunocompromised cohorts in 28 studies reporting data after the second dose.

Among immunocompromised groups, in incremental order, transplant recipients had the lowest pooled risk ratio of 0.06 (95%CI: 0.04 to 0.09, I^2=0%, p=0.81) (GRADE=Moderate) followed by haematological cancer patients at 0.36 (95%CI: 0.21 to 0.62, I^2 = 89%, p<0.01) (GRADE=Moderate), solid cancer patients at 0.40 (95%CI: 0.31 to 0.52, I^2 = 63%, p=0.03) (GRADE=Moderate) and IMID patients at 0.66 (95%CI: 0.48 to 0.91, I^2=81%, p<0.01) (GRADE=Moderate).

After the second dose, the lowest pooled risk ratio was again seen in transplant recipients at 0.29 (95%CI: 0.21 to 0.40, I^2=91%, p<0.01) (GRADE=Moderate), haematological cancer patients at 0.68 (95%CI: 0.57 to 0.80, I^2=68%, p=0.02) (GRADE=Low), IMID patients at 0.79 (95%CI: 0.72 to 0.86, I^2=87%, p<0.01) (GRADE=Low) and solid cancer at 0.92 (95%CI: 0.89 to 0.95, I^2=26%, p=0.25) (GRADE=Low).

**Conclusion:** Seroconversion rates and serological titres are significantly lower in immunocompromised patients with transplant recipients having the poorest outcomes. Additional strategies on top of the conventional 2-dose regimen will likely be warranted, such as a booster dose of the vaccine.

**Systematic review registration:** PROSPERO CRD42021272088

## INTRODUCTION

Spread of the severe acute respiratory syndrome coronavirus 2 (SARS-CoV-2) has led to the ongoing global COVID-19 pandemic. By the third quarter of 2021, there have been over 200 million confirmed cases and over 4 million deaths worldwide. The morbidity and mortality from COVID-19 and its complications and large-scale economic disruption have prompted an unprecedented pace in vaccine development.(1, 2) Vaccines which have been approved for use to date include new technology mRNA vaccines (e.g. Pfizer-BioNTech and Moderna), and non-replicating viral vector vaccines (e.g. Jannsen Ad26.COV2.S) and traditional inactivated vaccines (eg Sinovac).(3) Trials and ongoing studies have sought to evaluate the efficacy and safety of these vaccines. High vaccine efficacy against symptomatic laboratory-confirmed SARS-CoV-2 infection has been reported, with over 50% after the first dose and 90% after the second dose for the BioNTech-Pfizer vaccine(4), while Oxford-AstraZeneca reported an efficacy of 70% after the second dose.(5) High seroconversion rates were shown regardless of the vaccine received or previous infection status.(6)

However, vaccine trials have excluded immunocompromised groups, such as transplant recipients and patients with rheumatological conditions, leading to a paucity of data on the efficacy and safety of vaccines in these groups. These patients, which constitute about 3% of the adult population(7), are of particular interest due to possible suppression or over-activation of the immune system attributable to the primary disease or concurrent therapy. There is an urgent need for data and insights on this as infection and viral shedding have reported to be more severe and persistent.(8, 9) Patients with active cancer are recognized to be at increased risk of severe COVID-19 infection and death.(10, 11) Transplant recipients require prolonged immunosuppression to prevent the risk of graft rejection and past studies have shown increased risk of severe diseases and poor outcomes with COVID-19.(12) Patients with autoimmune and inflammatory rheumatic diseases requiring immunosuppressive treatment have worse outcomes from COVID-19 infection compared to age- and gender-comparable patients without such conditions.(13)

Past studies of other vaccines such as the influenza and pneumococcal vaccine among immunocompromised groups have shown variable efficacy depending on factors such as vaccine type, underlying disease and concurrent medications. In a meta-analysis on the immunogenicity of the influenza vaccination in organ transplant recipients, the risk factors of lower seroconversion included being within 6 months post-transplant, on anti-metabolites, and lung transplantation.(14) Other studies have shown reduced antibody response after the influenza vaccine among cancer patients, transplant recipients and those taking other anti-CD20 based immunosuppressive regimens like rituximab used in those with rheumatic conditions.(15, 16)

To date, there have been no systematic reviews looking at the immunogenicity of COVID-19 vaccines in immunocompromised cohorts. As such, this review aims to study seroconversion rates and antibody levels post-vaccination amongst immunocompromised patients compared to healthy controls.

## METHODOLOGY

The systematic review is reported according to the Preferred Reporting Items for Systematic Reviews and Meta-Analyses (PRISMA) guidelines (Supplementary table 1).(17) This review is registered with the National Institute for Health Research international prospective register of systematic reviews (PROSPERO) at CRD42021272088.

### Search strategy

Searches of databases MEDLINE via PubMed, EMBASE, Cochrane Central Register of Controlled Trials (CENTRAL), CORD-19, WHO COVID-19 Research Database, ClinicalTrials.gov and WHO international clinical trials registry platform were conducted per protocol in September 2021 for articles published from 1 December 2020 to 3 September 2021. There was no restriction on language of publication. Literature search was performed using the search strategy in each database in Table 1. To improve validity of data, non-peer-reviewed preprints from preprint databases were not used.

**Table 1:** Search strategy.

| Database | Search terms | Results |
| --- | --- | --- |
| CENTRAL | ("coronavirus" OR "coronavirus" OR "covid 2019" OR "SARS2" OR "SARS-CoV-2" OR "SARS-CoV-19" OR "novel cov" OR "2019ncov" OR "sars cov2" OR "cov22" OR "ncov" OR "covid-19" OR "covid19" OR "coronaviridae" OR "corona virus") in All Fields | 3,788 |
|  | AND ("vaccine" OR "vaccination" OR vaccine OR vaccination OR vaccin*) in All Fields | 1,563 |
|  | AND ("cancer" OR "cancer" OR "malignancy" OR "malign*" OR "immunocompromise" OR "immunocompromised" OR "immunodeficiency" OR "immunodeficient" OR "immunodef*" OR "immunocompr*" OR "chemotherapy" OR "chemo*" OR "immunosuppressed" OR "immunosuppression" OR "immunosupp*" OR "rheumatology" OR "rheumatic" OR "rheum*" OR "autoimmune" OR "autoimmunity" OR "transplant" OR "solid organ" OR steroids OR antineoplastic agents OR chemotherapy OR cytotoxicity OR immunologic OR antirheumatic agents OR immunosuppressive agents or steroid* or corticosteroid* or (antineoplastic* AND agent*) OR chemotherap* or cytotoxic*) in All Fields | 17,945 |
|  | All filters activated | 152 |
|  | Search limits: 1 December 2020 to 3 September 2021 |  |
| MedLine via PubMed | ("coronavirus"[MeSH] OR "coronavirus"[All Fields] OR "covid 2019"[All Fields] OR "SARS2"[All Fields] OR "SARS-CoV-2"[All Fields] OR "SARS-CoV-19"[All Fields] OR "novel cov"[All Fields] OR "2019ncov"[All Fields] OR "sars cov2"[All Fields] OR "cov22"[All Fields] OR "ncov"[All Fields] OR "covid-19"[All Fields] OR "covid19"[All Fields] OR "coronaviridae"[All Fields] OR "corona virus"[All Fields])<br>AND<br>("vaccine"[MeSH] OR "vaccination"[MeSH] OR vaccine[All Fields] OR vaccination[All Fields] OR vaccin*[All Fields])<br>AND<br>("cancer"[MeSH] OR "cancer" OR "malignancy" OR "malign*" OR "immunocompromise"[All Fields] OR "immunocompromised"[All Fields] OR "immunodeficiency"[All Fields] OR "immunodeficient"[All Fields] OR "immunodef*" [All Fields] OR "immunocompr*" [All Fields] OR "chemotherapy"[All Fields] OR "chemo*" [All Fields] OR "immunosuppressed"[All Fields] OR "immunosuppression"[All Fields] OR "immunosupp*" [All Fields] OR "rheumatology"[All Fields] OR "rheumatic"[All Fields] OR "rheum*" [All Fields] OR "autoimmune"[All Fields] OR "autoimmunity"[All Fields] OR "transplant"[All Fields] OR "solid organ"[All Fields] OR steroids[MeSH] OR antineoplastic agents[MeSH] OR chemotherapy[MeSH] OR cytotoxicity[MeSH] OR immunologic[MeSH] OR antirheumatic agents[MeSH] OR immunosuppressive agents[MeSH] or steroid* or corticosteroid* or (antineoplastic* AND agent*) OR chemotherap* or cytotoxic*)<br>Search limits: 1 December 2020 to 3 September 2021 | 2,571 |
| EMBASE | ('coronavirinae'/exp OR 'coronavirinae' OR 'coronaviridae infection'/exp OR 'coronaviridae infection' OR 'coronavirus disease 2019'/exp OR 'coronavirus'/exp OR coronavirus OR 'coronavirus infection'/de) NOT [medline]/lim<br>AND<br>('vaccination'/exp OR vaccine OR vaccination OR vaccin*)<br>AND<br>("cancer"/exp OR "malignancy"/exp OR "malign*" OR "immunocompromise"/exp OR "immunocompromised" OR "immunodeficiency"/exp OR "immunodeficient" OR "immunodef*" OR "immunocompr*" OR "chemotherapy"/exp OR "chemo*" OR "immunosuppressed"/exp OR "immunosuppression" OR "immunosupp*" OR "rheumatology"/exp OR "rheumatic" OR "rheum*" OR "autoimmune"/exp OR "autoimmunity" OR "transplant"/exp OR "solid organ"/exp)<br>Search limits: 1 December 2020 to 3 September 2021 | 724 |

### Study selection

A two-staged screening method was adopted, screening by title and abstract and screening by full-text article. Each title, abstract and full-text identified was screened independently by two researchers with discrepancies resolved by consultation of a third researcher. Results were limited to human subjects. Studies of any follow-up duration and timepoints were included.

We included published and unpublished prospective observational and experimental studies that met the following criteria:

- Studies that involved human participants all of whom should be receiving a COVID- 19 vaccine of any brand and type
- Studies that involved patients with solid organ malignancies, haematological malignancies, organ transplant recipients and patients with immune-mediated inflammatory disorders (IMIDs)
- Studies that included and reported data of a control group comprising healthy individuals or comparators who are not immunocompromised defined as not having malignancy, rheumatologic, autoimmune and organ transplant conditions
- Studies that reported at least one of the following outcome measures:

- Seroconversion after COVID-19 vaccination
- Serological titres after COVID-19 vaccination

Studies not adhering to the aforementioned inclusion criteria were excluded. Additionally, studies were excluded if they:

- Included but did not report outcomes of an immunocompetent control group
- Reported seroconversion data in a form from which proportions, risk of seroconversion or number of seroconverted participants could not be derived
- Reported serological titres in a form from which neither mean nor median titres could be derived

### Data extraction

Data was extracted according to a pre-determined proforma in Microsoft Excel Version 16.45 by two researchers. All key extracted data was reviewed and quality-checked at the end of the data-extraction phase.

Study characteristics comprised setting, primary and secondary outcomes, study design, sample size, dropout and non-response rates and inclusion and exclusion criteria. Participant data collected comprised age, sex and comprehensive disease and treatment history, including immunosuppressive regimen. Intervention-related data included vaccine type and brand, dosing schedule and number of subjects receiving each type and brand of vaccine and median or mean interval between doses. Outcome-related data comprised assay, antibody measured and method of measurement, intervals of sample collection and number of measurements made.

### Risk of Bias assessment

The Risk Of Bias In Non-randomized Studies of Interventions (ROBINS-I) tool was used to rate risk of bias for included non-randomised studies which assesses 7 domains: Risk of bias due to 1) confounding 2) selection of participants into the study 3) classification of interventions 4) deviations from intended interventions 5) missing data 6) measurement of outcomes 7) selection of the reported results.(18)

The Cochrane Risk of Bias 2.0 tool was planned to be used for experimental studies which assesses 5 domains: bias arising from 1) the randomisation process, 2) deviations from intended interventions, 3) missing outcome data, 4) measurement of the outcome and 5) bias in selection of the reported result. However, no experimental studies were yielded in our search.(19)

Two reviewers assessed each paper in parallel and reached consensus by discussion. All discrepancies were resolved by involving a third reviewer assessing the paper independently.

### Data analysis

Our search did not identify any randomised trials involving the use of COVID-19 vaccines in immunocompromised patients. We performed a meta-analysis of associations by pooling risk ratios (RRs) from observational studies using DerSimonian random effects meta-analysis. Sensitivity analysis was performed by comparing the results to other meta-analysis models including fixed effect models and Knapp-Hartung random effects models, and excluding trials with high risk of bias. Publication bias was assessed visually using funnel plots. Subgroup analysis and mixed-effects meta-regression was conducted according to average age, vaccine type, risk of bias, timepoints, brand of serology kit and country of study. The synthesis without meta-analysis approach was used to summarize the data qualitatively when meta-analysis of the data is not feasible due to variation in the reporting of outcomes of interests. All analyses were run using R Version 4.1.0.

### Certainty of evidence

We assessed the certainty of evidence using the Grading of Recommendations Assessment, Development and Evaluation (GRADE).(20) Certainty of evidence for each study was rated as high, moderate, low, or very low, based on considerations of risk of bias, inconsistency, indirectness, publication bias, intransitivity, incoherence and imprecision.

### Patient and public involvement

No patients or members of the public were directly involved in this research study.

## RESULTS

The results of our screening is illustrated in Figure 1. We identified 42 studies for our systematic review.(21–62) Table 2 outlines the details of all included studies.

**Figure 1:**
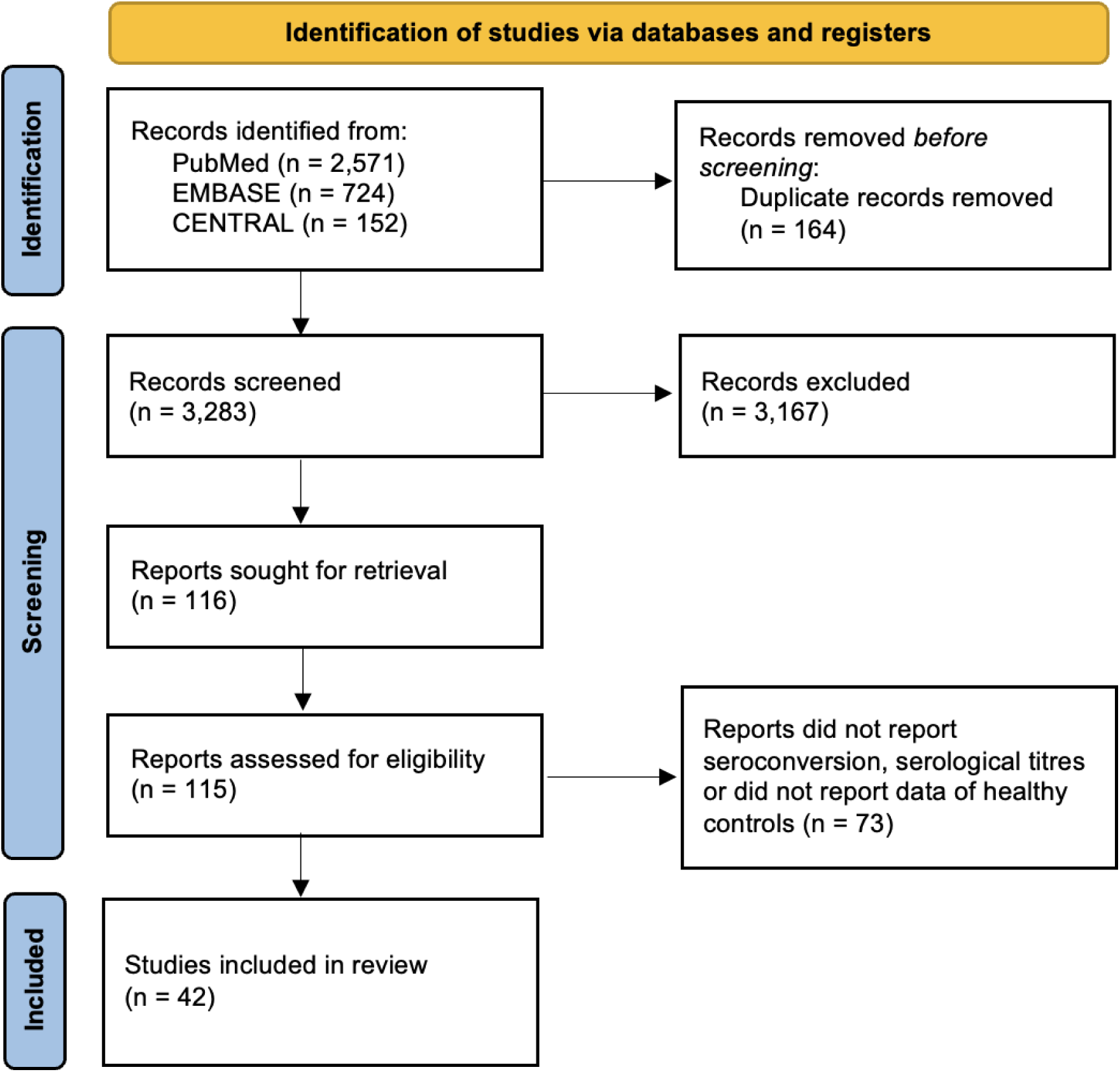
PRISMA flowchart

**Table 2:**
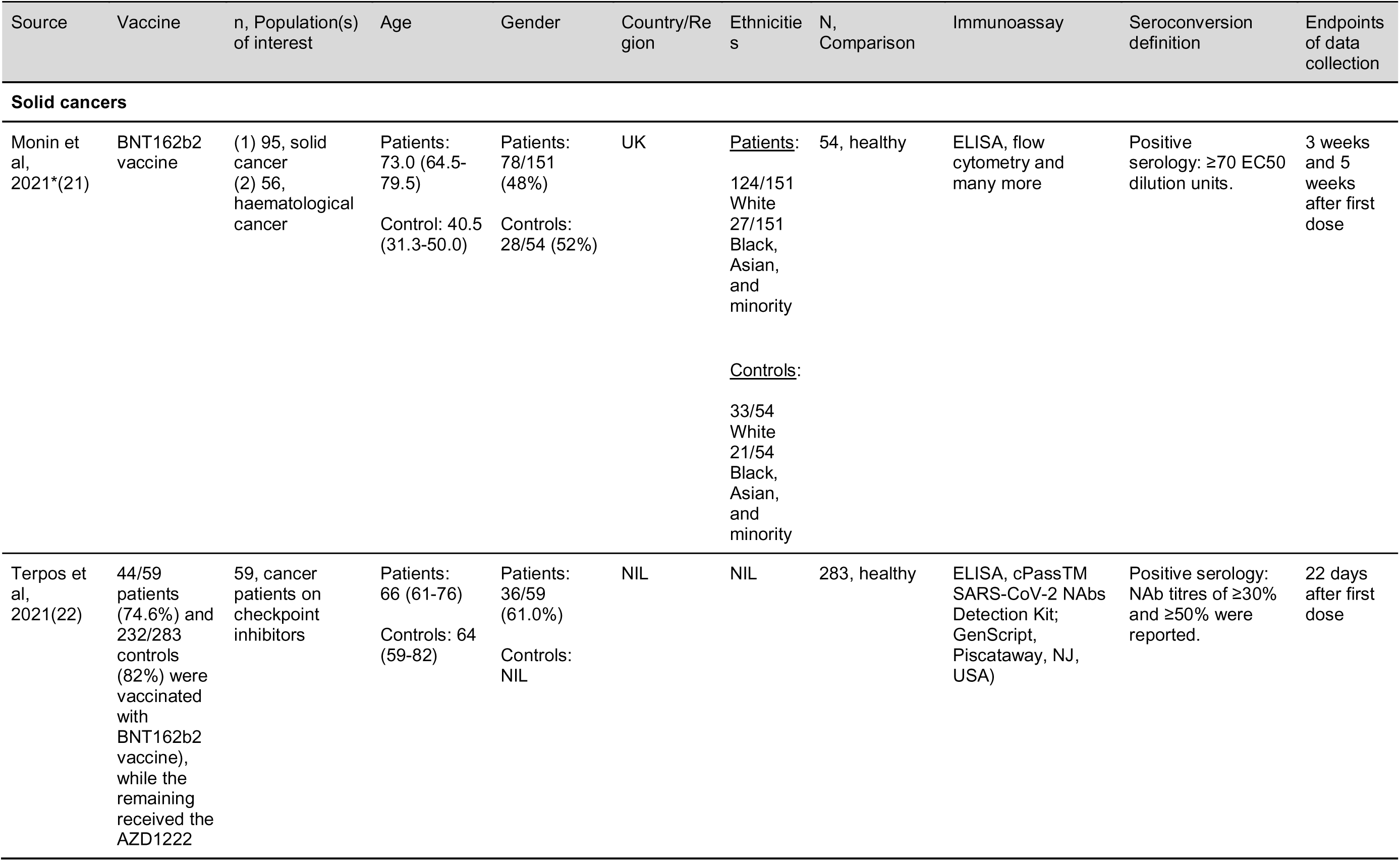
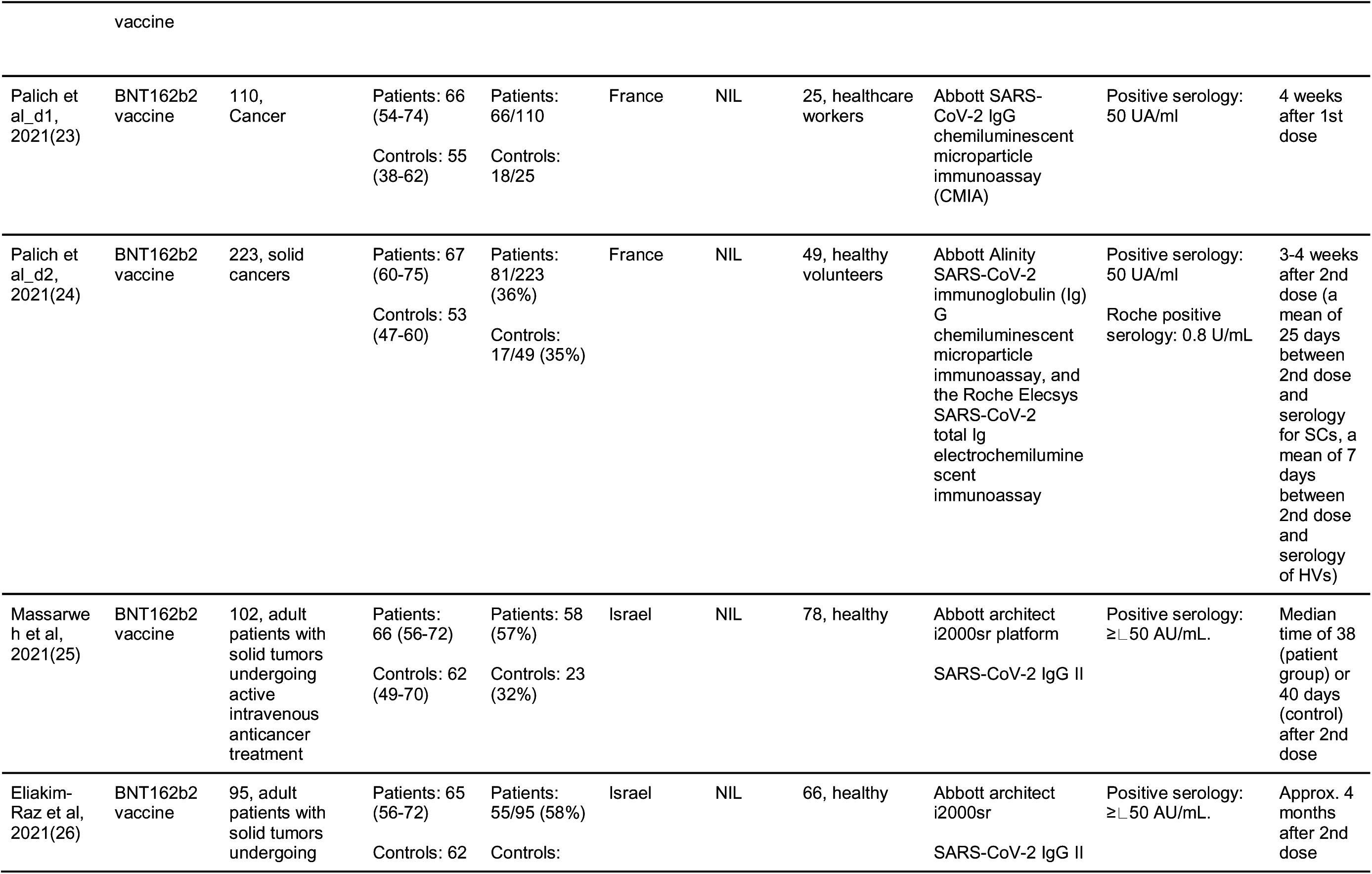
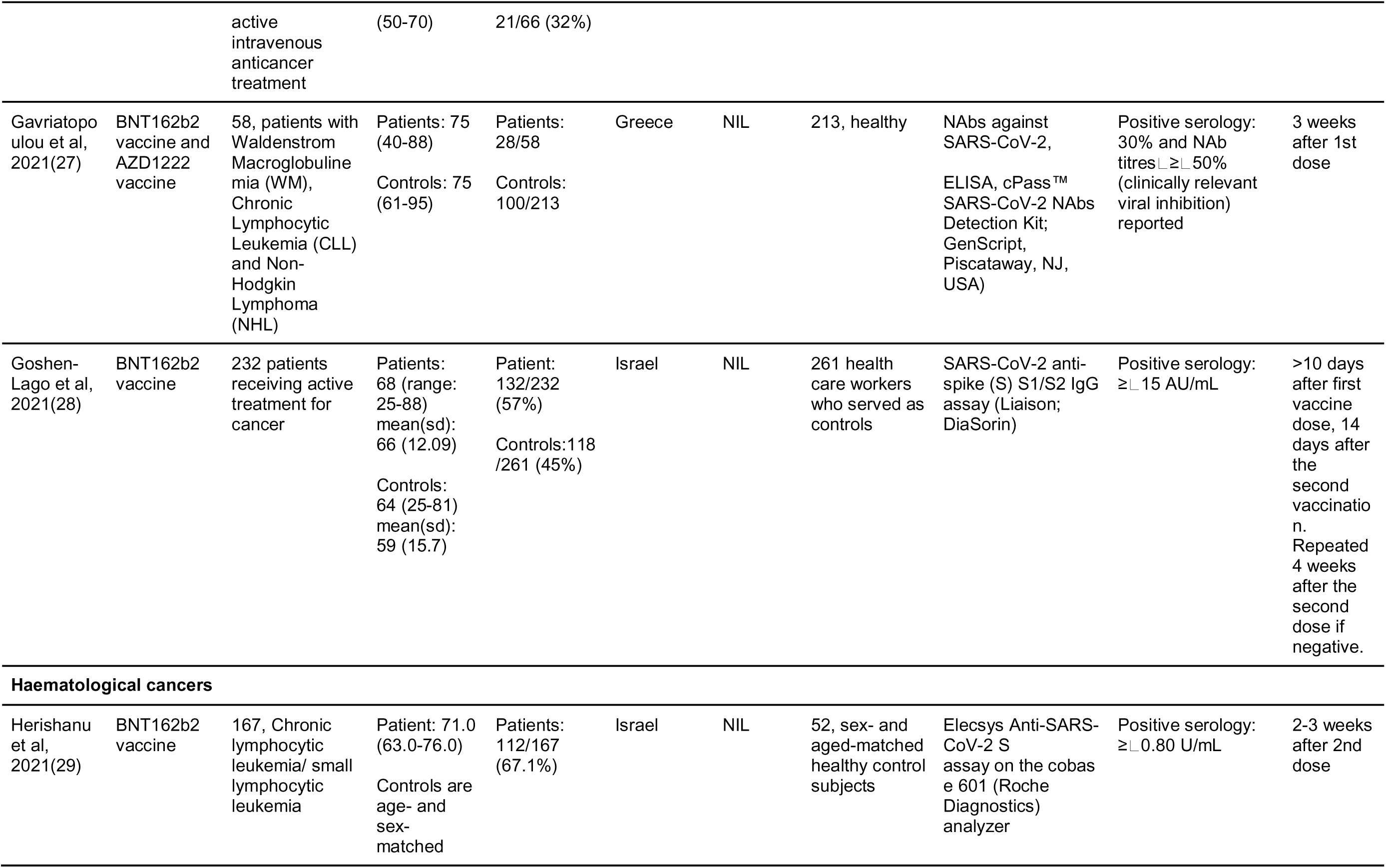
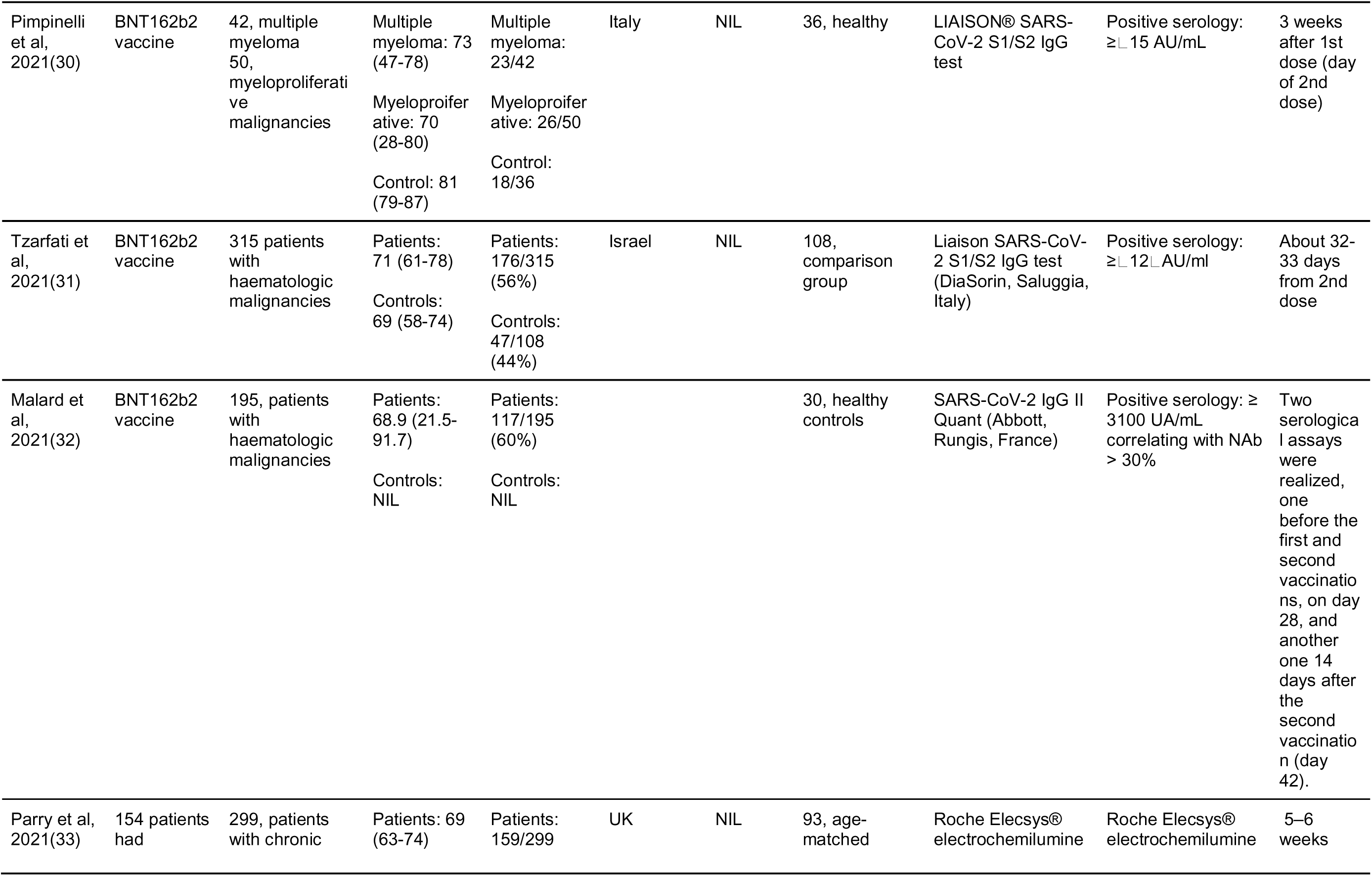
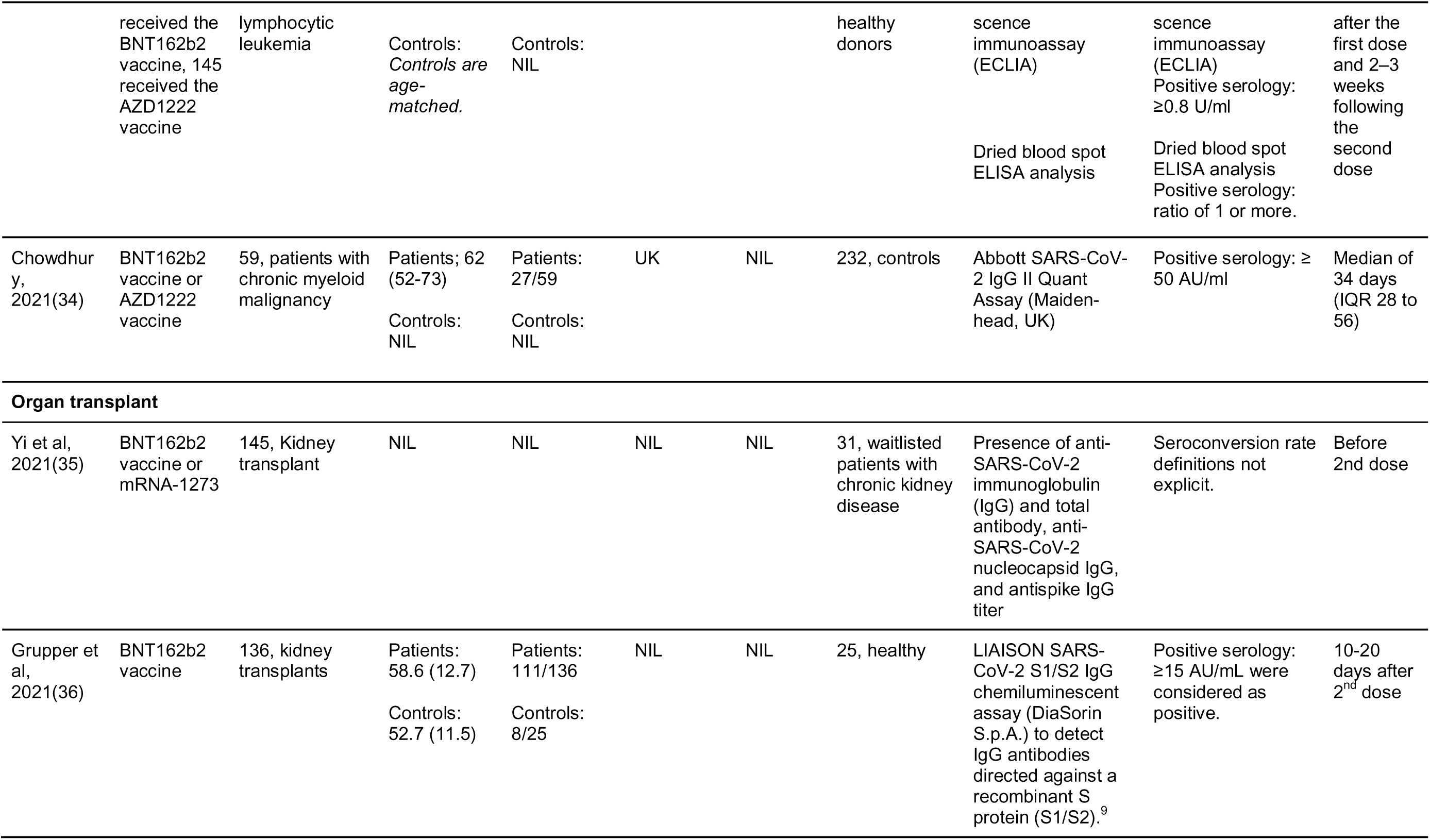
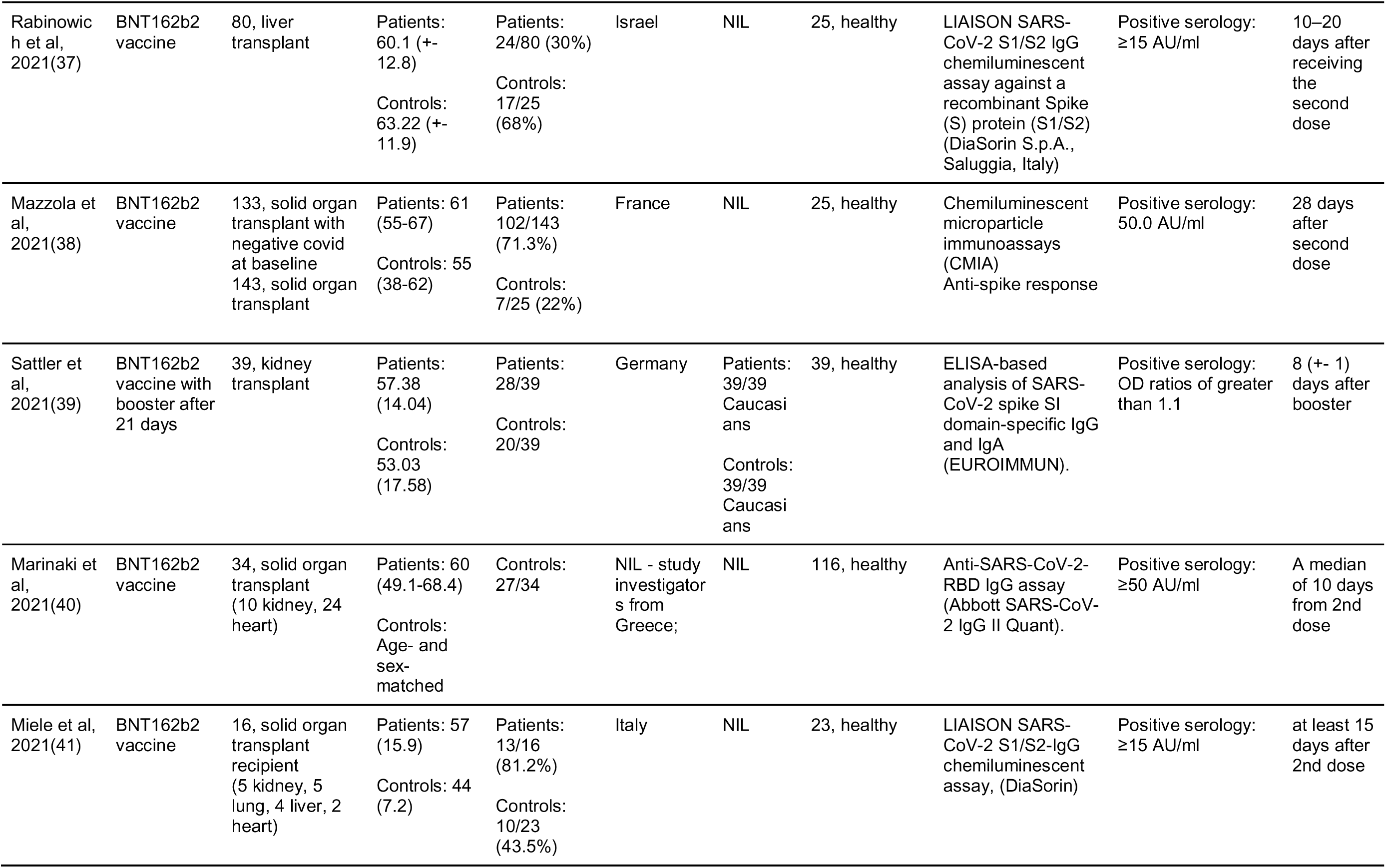
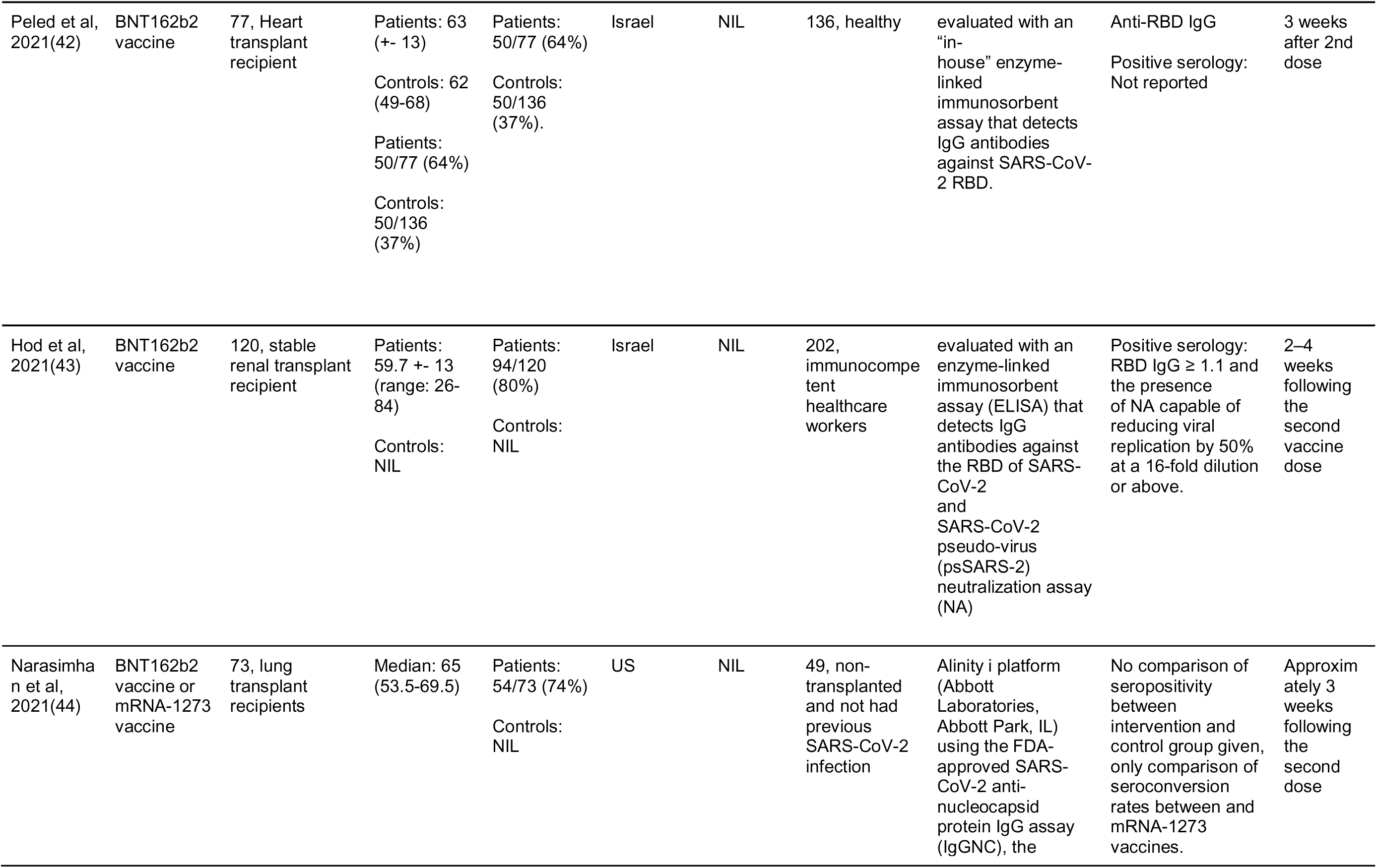
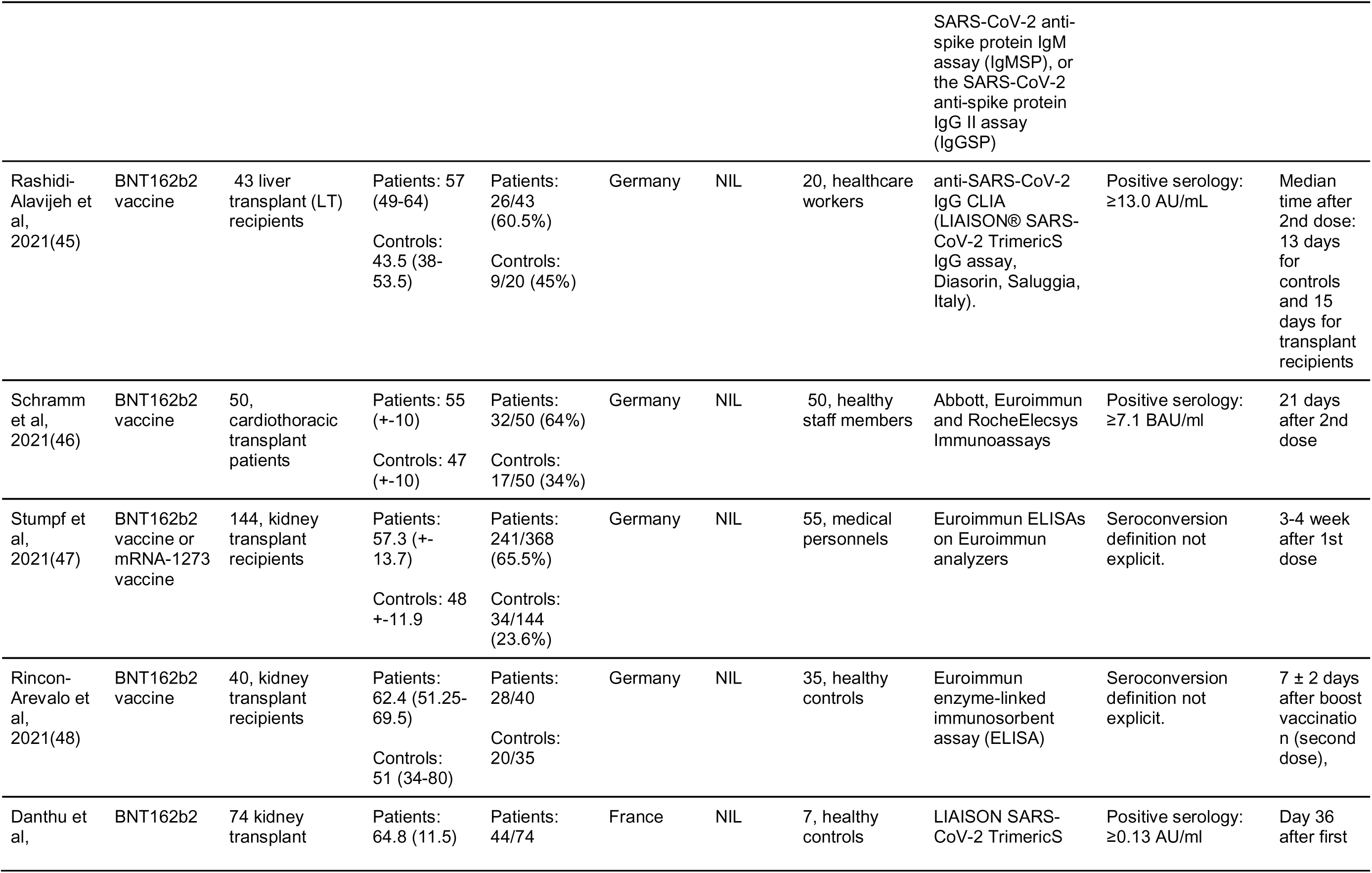
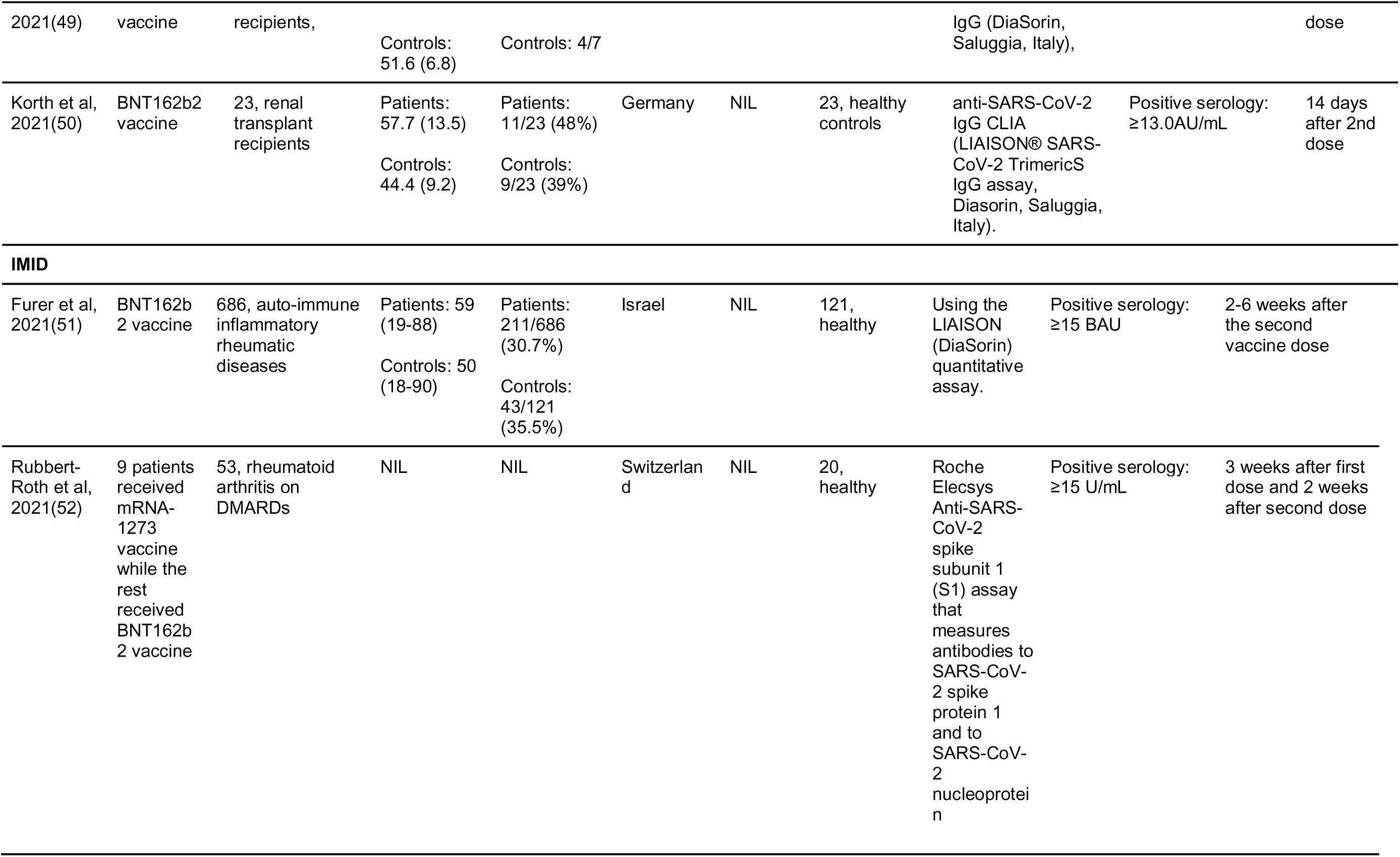
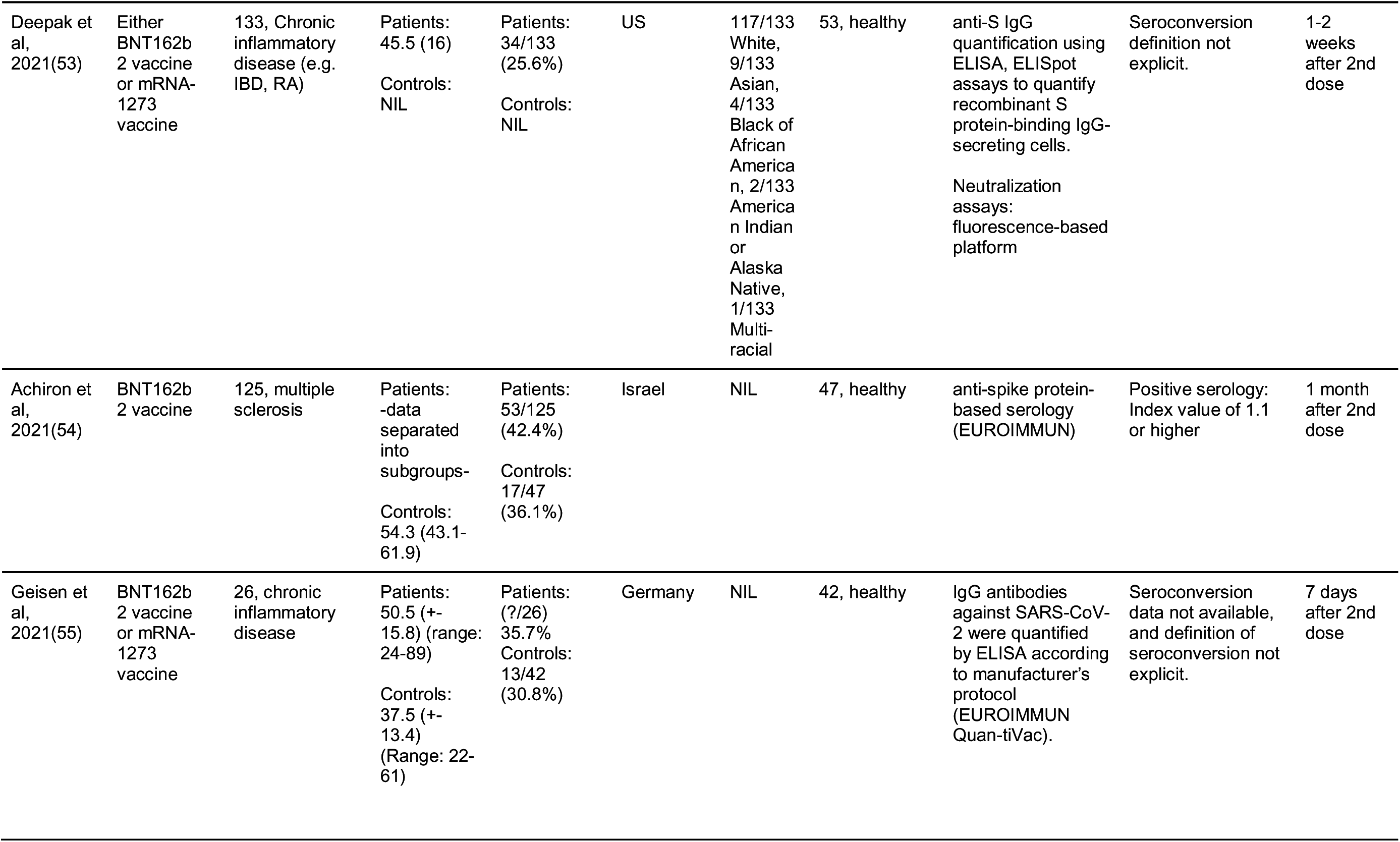
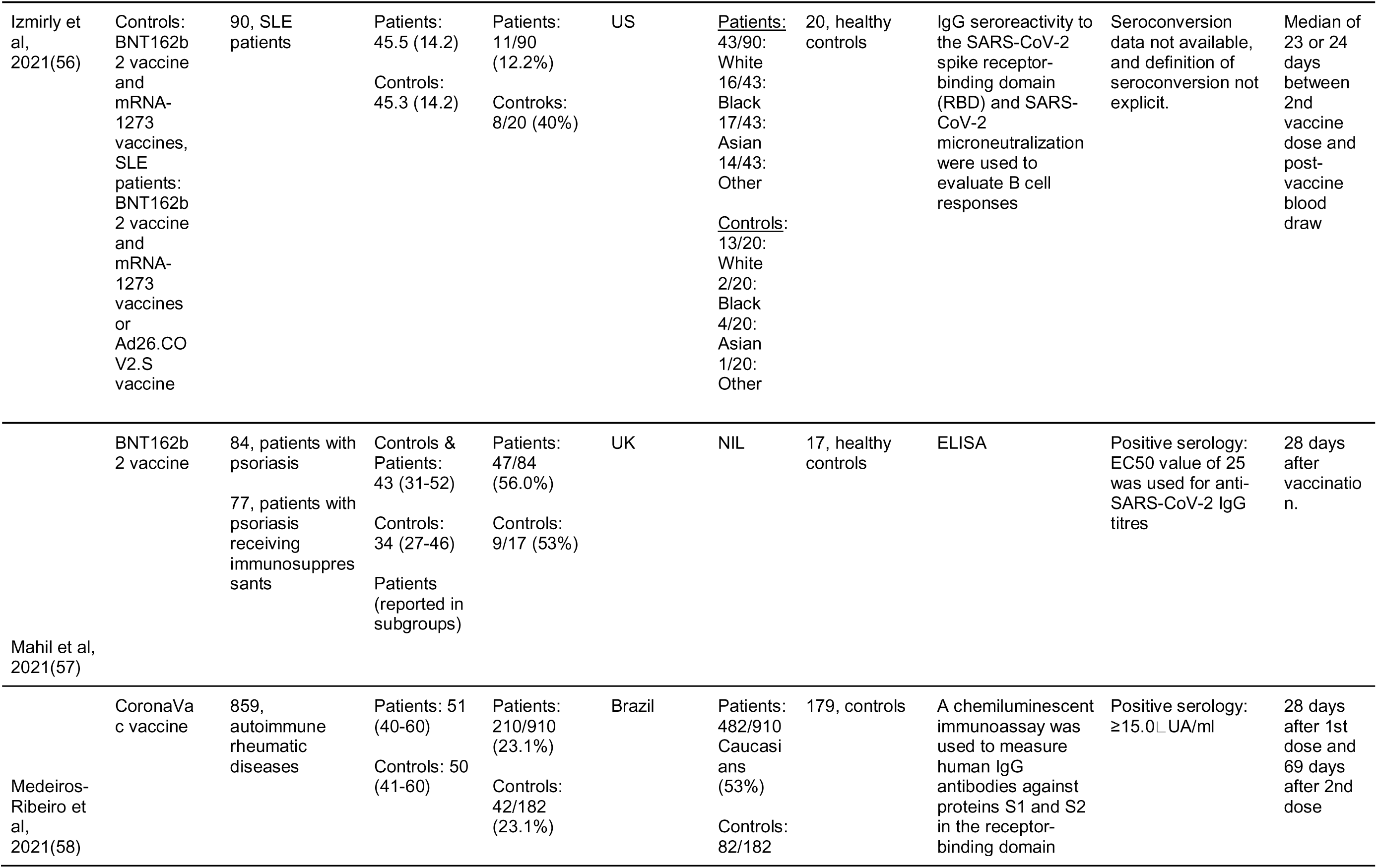
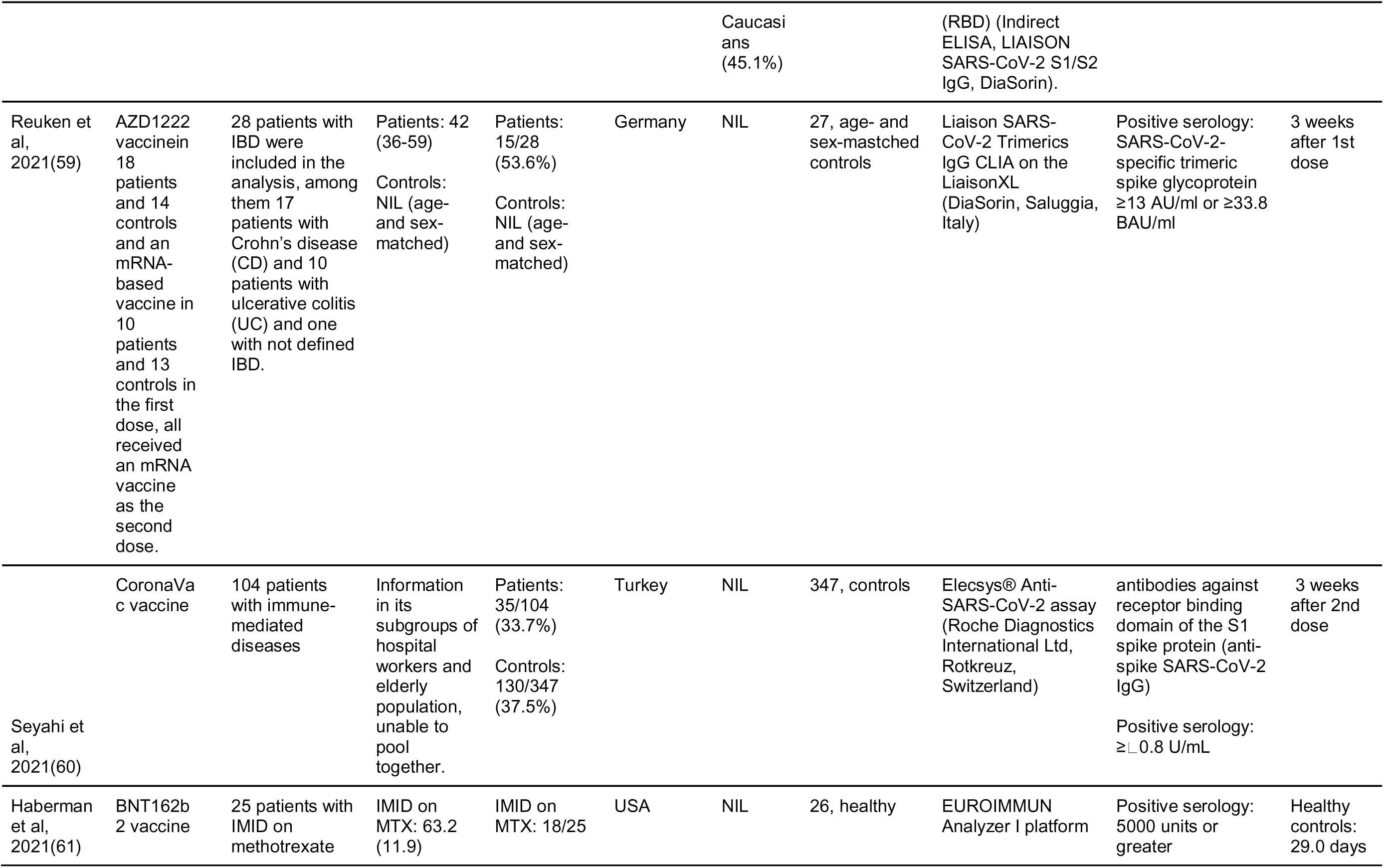
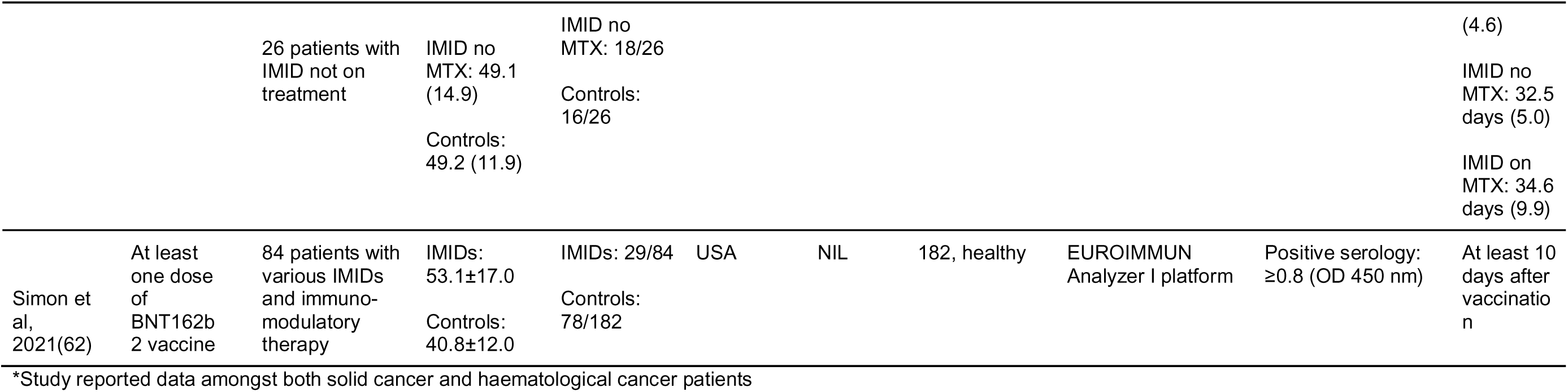
Trial characteristics.

Studies were all observational in nature with no experimental trials identified relevant to our study population. Only prospective comparative studies were included for this meta-analysis. Case reports, series, qualitative studies and retrospective studies were excluded to better inform estimates of effect.

We analysed seroconversion rates and antibody titre levels. Direct evidence of vaccine protection in immunocompromised patients was not used. Vaccines administered include BNT162b2 (Pfizer-BioNTech), mRNA-1273 (Moderna), AZD1222 (AstraZeneca), Ad26.COV2.S (Janssen) and CoronaVac (Sinovac Biotech).

At the time where the studies were conducted, recommended vaccine regimens were 2-dose regimens. This meta-analysis further stratifies the results according to post-first dose and post-second dose seroconversion and antibody levels.

In summary, trials primarily included immunocompromised groups of cancer, organ transplant and IMID patients. 8/42 studies involving solid cancer patients, 7/42 studies involving haematological cancer patients, 12/42 studies involving IMID patients, and 16/42 studies involving organ transplant recipients.

### Outcomes of interest

The primary outcomes of interest were seroconversion after 1st and 2nd doses of COVID-19 vaccination. As brand and type of assay, immunoglobulin and definition of seroconversion differed across studies, the respective data was extracted from each study and reported in Table 2.

Secondary outcomes of interest were mean or median serological titres after 1st and 2nd doses of COVID-19 vaccination. Similarly, as specific antibodies measured and reported differed across studies, the antibody measured was reported in Tables 3 and 4.

**Table 3:**
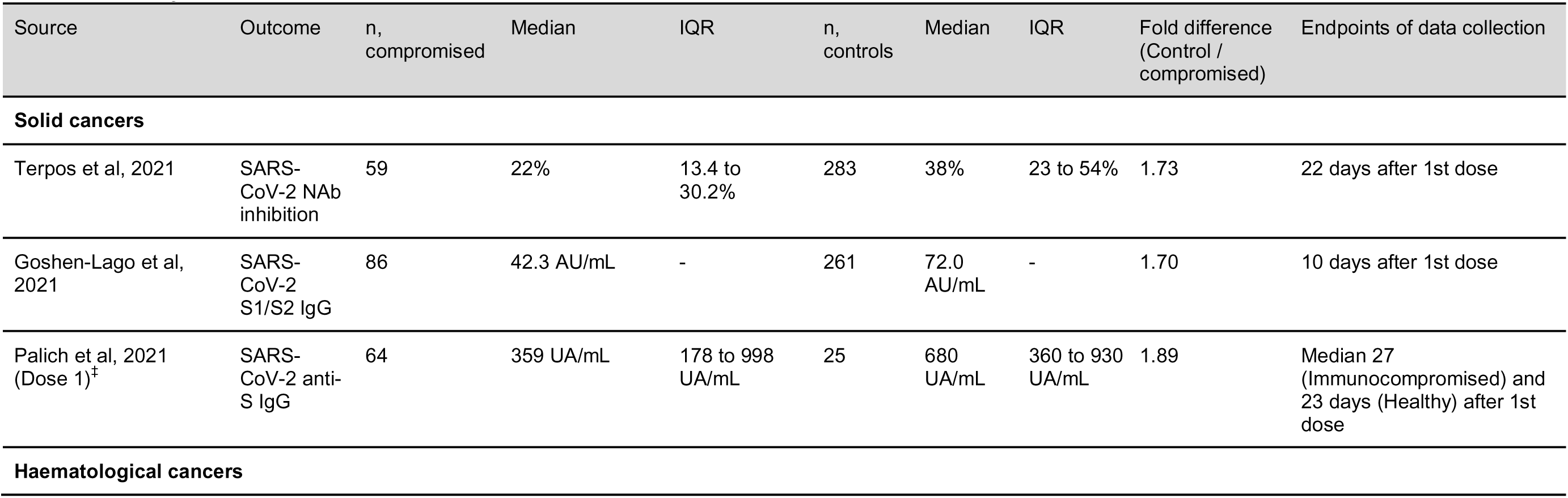
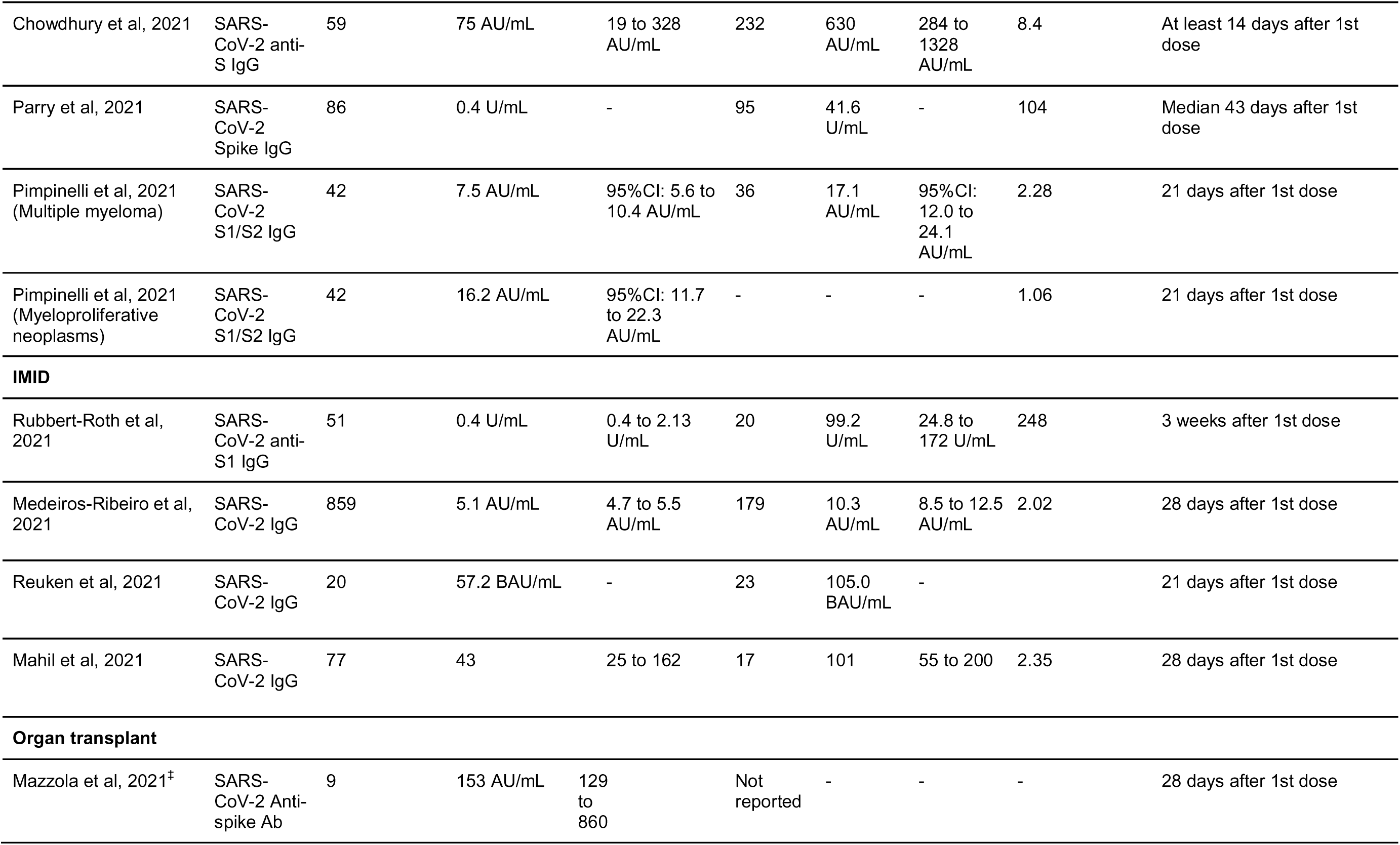
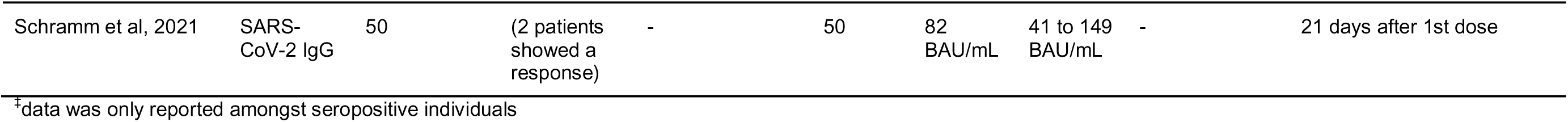
Antibody titres after the first dose of COVID-19 vaccine.

|  |  |  |  |  |  |  |  |  |  |
| --- | --- | --- | --- | --- | --- | --- | --- | --- | --- |
| Chowdhury et al, 2021 | SARS-CoV-2 anti-S IgG | 59 | 75 AU/mL | 19 to 328 AU/mL | 232 | 630 AU/mL | 284 to 1328 AU/mL | 8.4 | At least 14 days after 1st dose |
| Parry et al, 2021 | SARS-CoV-2 Spike IgG | 86 | 0.4 U/mL | - | 95 | 41.6 U/mL | - | 104 | Median 43 days after 1st dose |
| Pimpinelli et al, 2021 (Multiple myeloma) | SARS-CoV-2 S1/S2 IgG | 42 | 7.5 AU/mL | 95%CI: 5.6 to 10.4 AU/mL | 36 | 17.1 AU/mL | 95%CI: 12.0 to 24.1 AU/mL | 2.28 | 21 days after 1st dose |
| Pimpinelli et al, 2021 (Myeloproliferative neoplasms) | SARS-CoV-2 S1/S2 IgG | 42 | 16.2 AU/mL | 95%CI: 11.7 to 22.3 AU/mL | - | - | - | 1.06 | 21 days after 1st dose |
| **IMiD** |  |  |  |  |  |  |  |  |  |
| Rubbert-Roth et al, 2021 | SARS-CoV-2 anti-S1 IgG | 51 | 0.4 U/mL | 0.4 to 2.13 U/mL | 20 | 99.2 U/mL | 24.8 to 172 U/mL | 248 | 3 weeks after 1st dose |
| Medeiros-Ribeiro et al, 2021 | SARS-CoV-2 IgG | 859 | 5.1 AU/mL | 4.7 to 5.5 AU/mL | 179 | 10.3 AU/mL | 8.5 to 12.5 AU/mL | 2.02 | 28 days after 1st dose |
| Reuken et al, 2021 | SARS-CoV-2 IgG | 20 | 57.2 BAU/mL | - | 23 | 105.0 BAU/mL | - | - | 21 days after 1st dose |
| Mahil et al, 2021 | SARS-CoV-2 IgG | 77 | 43 | 25 to 162 | 17 | 101 | 55 to 200 | 2.35 | 28 days after 1st dose |
| **Organ transplant** |  |  |  |  |  |  |  |  |  |
| Mazzola et al, 2021<sup>†</sup> | SARS-CoV-2 Anti-spike Ab | 9 | 153 AU/mL | 129 to 860 | Not reported | - | - | - | 28 days after 1st dose |
| Schramm et al, 2021 | SARS-CoV-2 IgG | 50 | (2 patients showed a response) | - | 50 | 82 BAU/mL | 41 to 149 BAU/mL | - | 21 days after 1st dose |

\*data was only reported amongst seropositive individuals

**Table 4:**
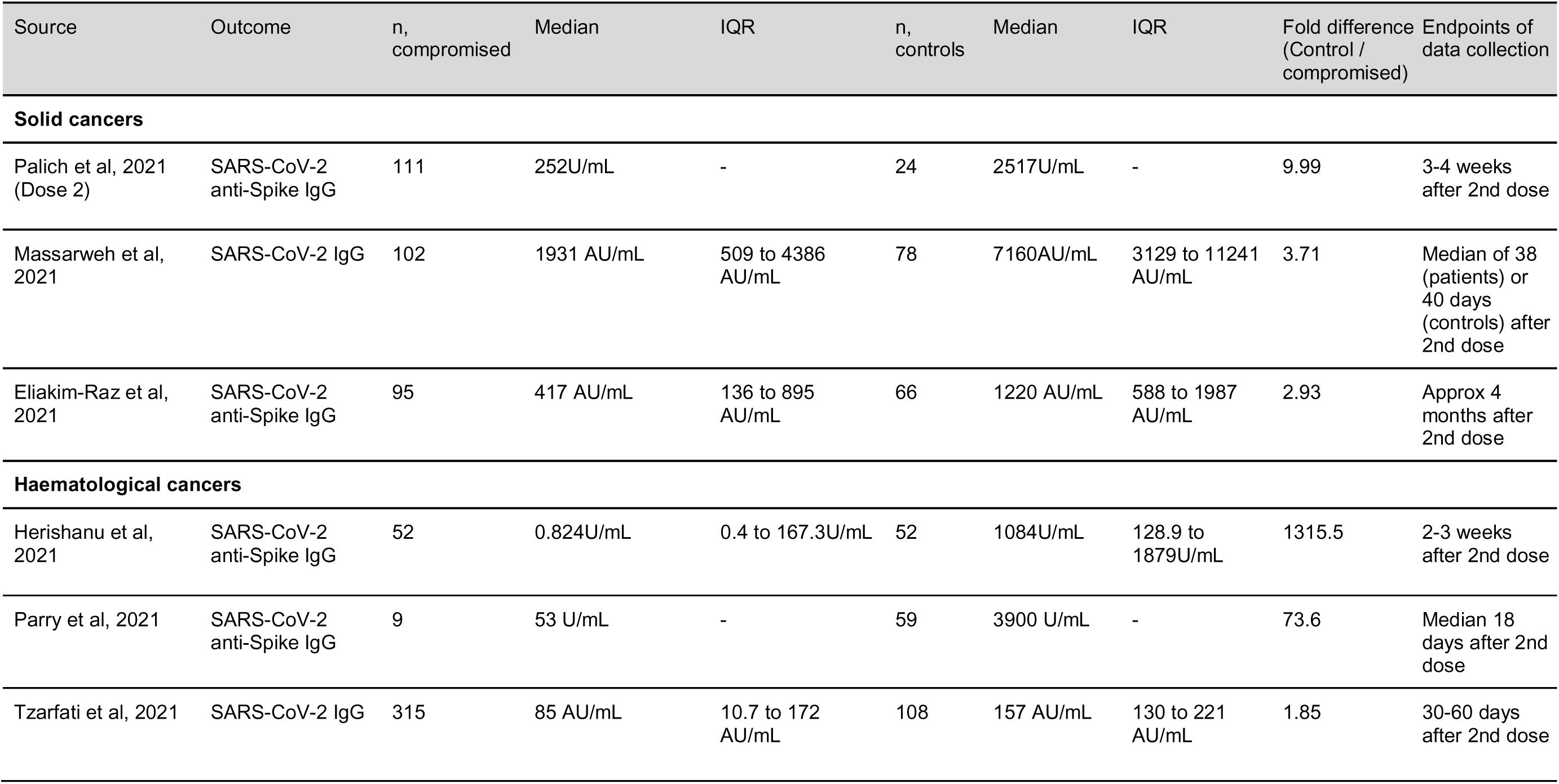
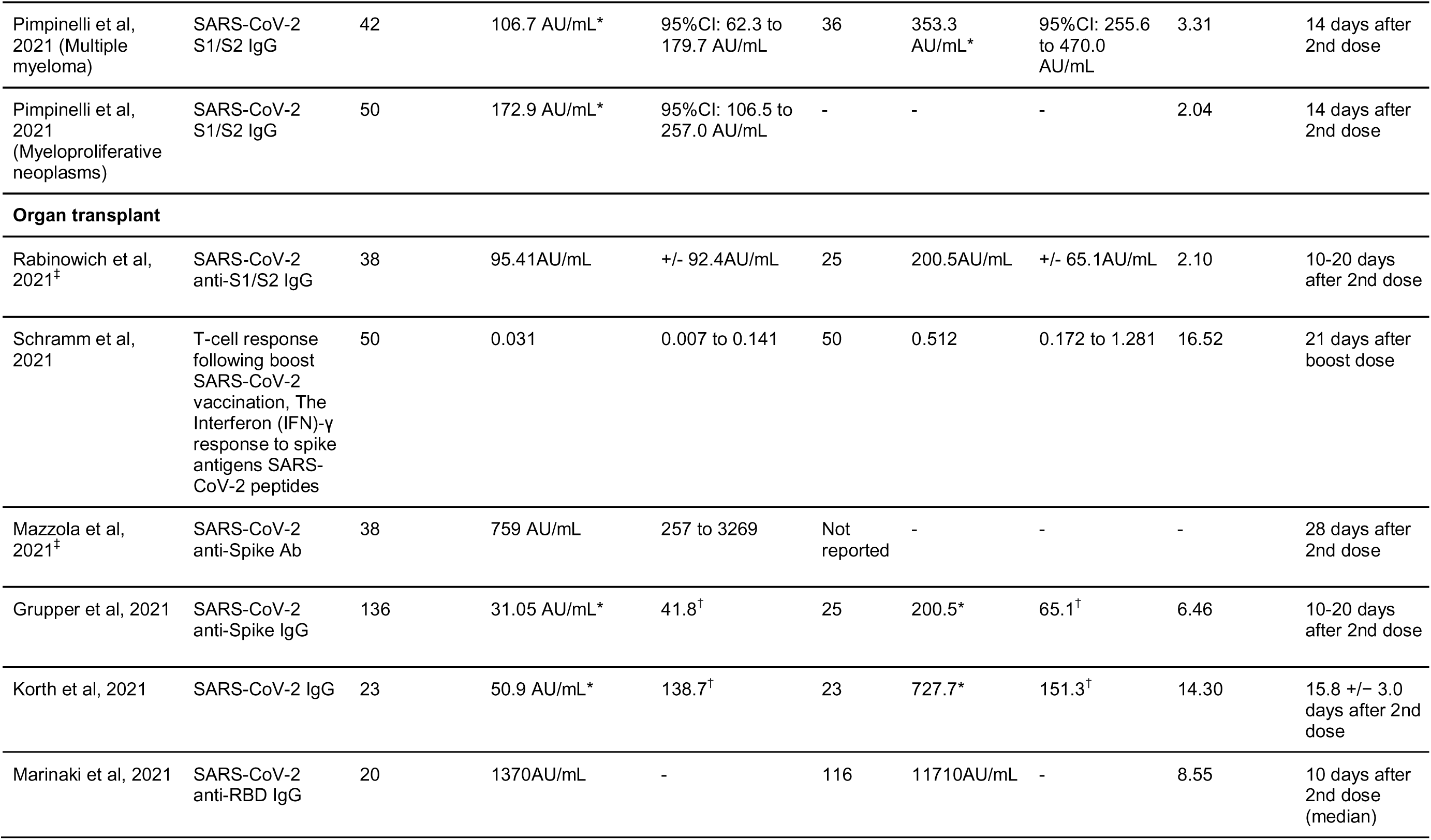
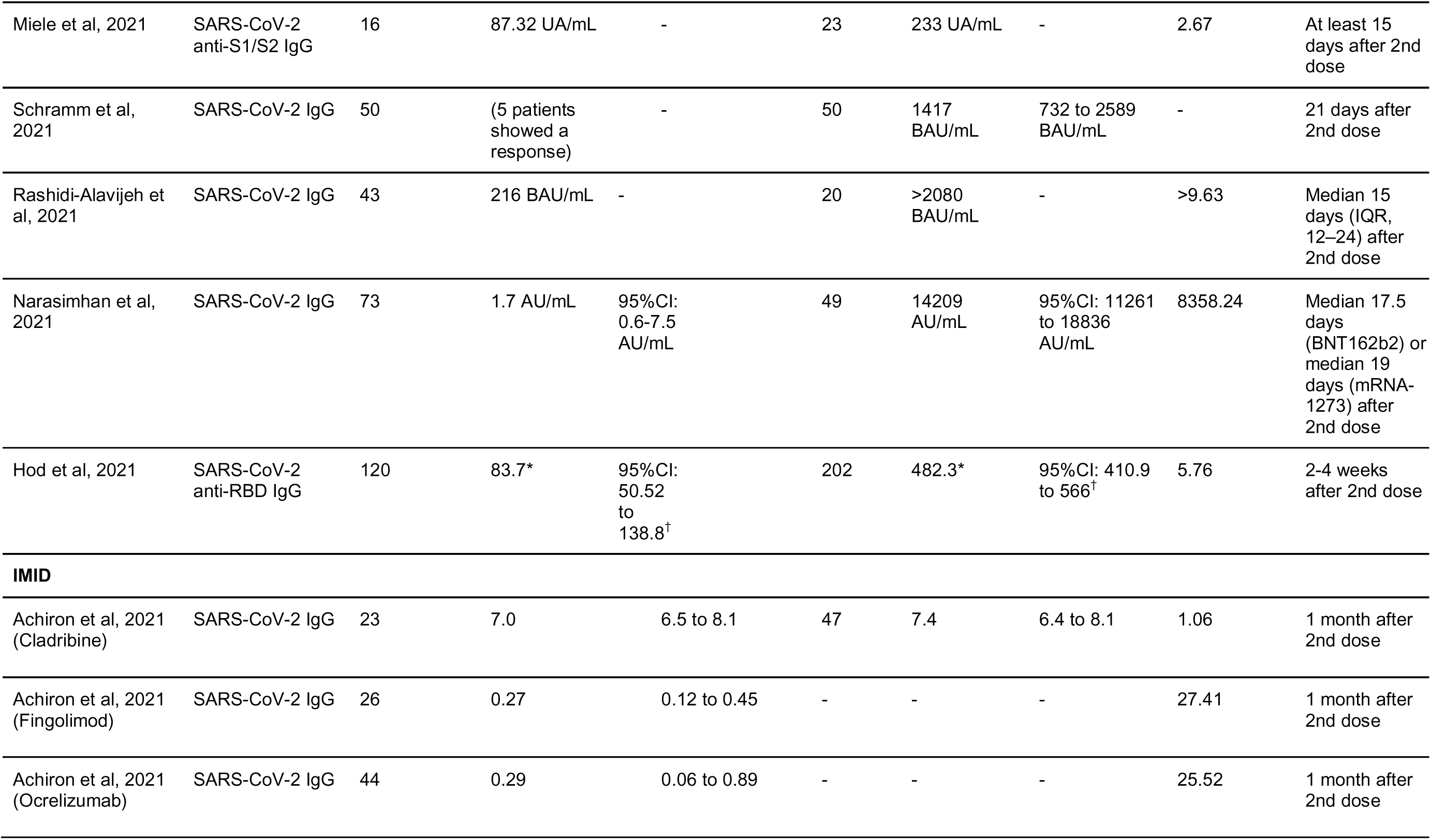
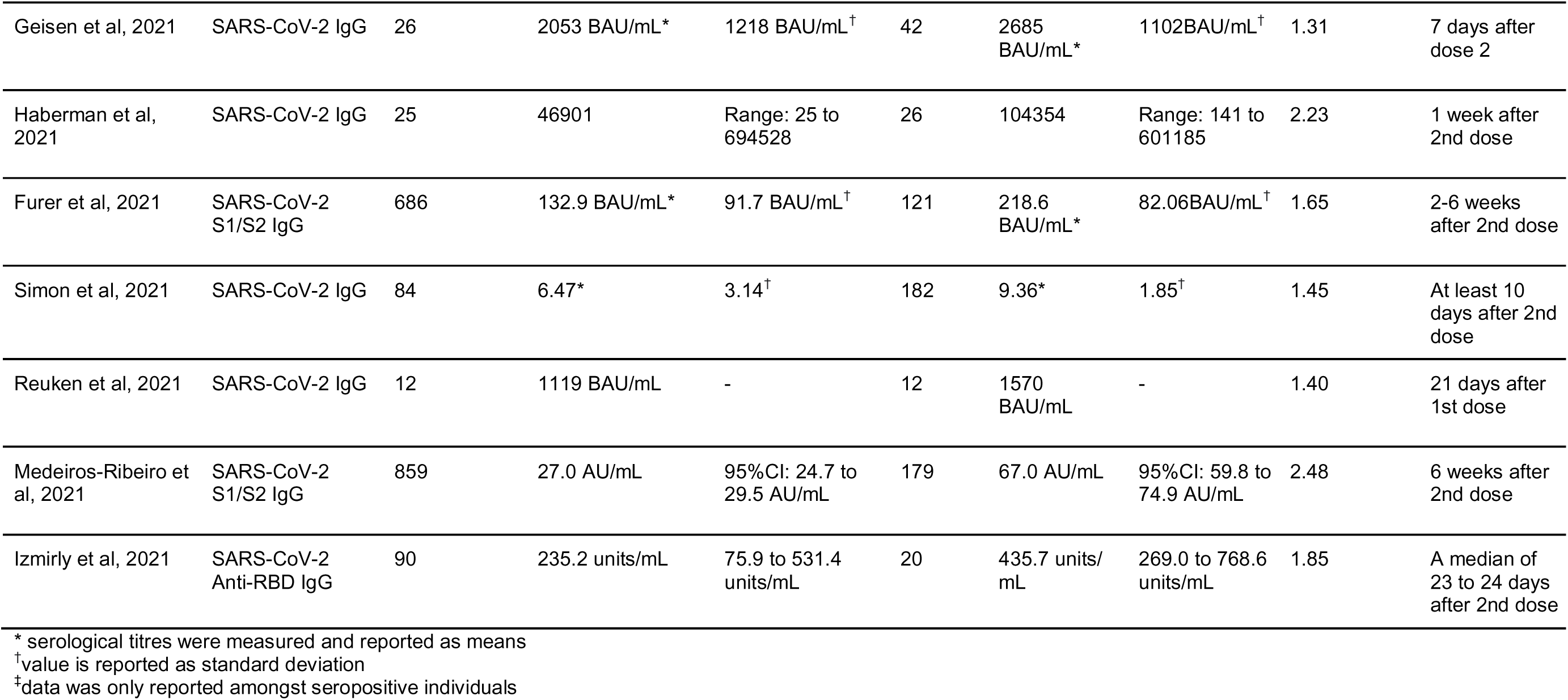
Antibody titres after a second dose of COVID-19 vaccine.

37/42 (88.0%) studies reported seroconversion rates in the immunocompromised and control groups. 16 studies (38.1%) assessed serological responses after the first dose of vaccine. 28/42 (66.7%) studies assessed the serological responses after the second dose of vaccine. The timepoints after COVID-19 vaccine of serological assessment and the different brands of serological kits used were extracted and reported in Table 2.

Solely mRNA vaccines were used in 35 (83.3%) of the studies, namely the BNT162b2 (Pfizer-BioNTech) and mRNA-1273 (Moderna) vaccine, 2 studies (4.8%) used the inactivated CoronaVac (Sinovac BioTech) vaccine and 5 studies (11.9%) involved the use of both mRNA and non-replicating viral vector, AZD1222 (AstraZeneca) or Ad26.COV2.S (Janssen), vaccines.

## SYNTHESIS OF RESULTS

### Dose 1

Figure 2 and Figure 3 respectively present the proportion with seroconversion and risk ratio of seroconversion amongst immunocompromised patients and healthy controls after the first dose of COVID-19 vaccine.

**Figure 2:**
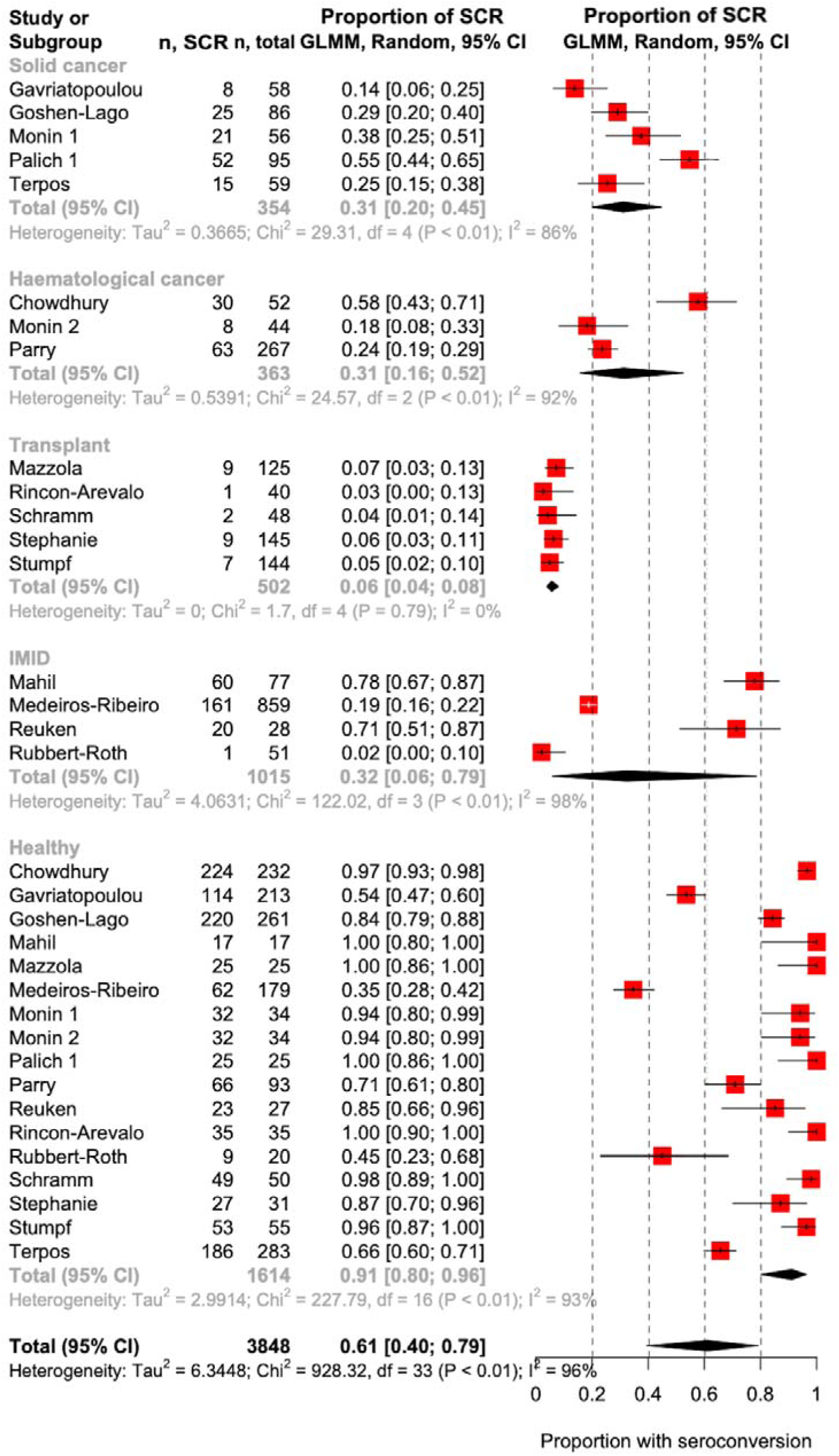
Proportion with seroconversion after the first dose of COVID-19 vaccine; SCR: Seroconversion after first dose

**Figure 3:**
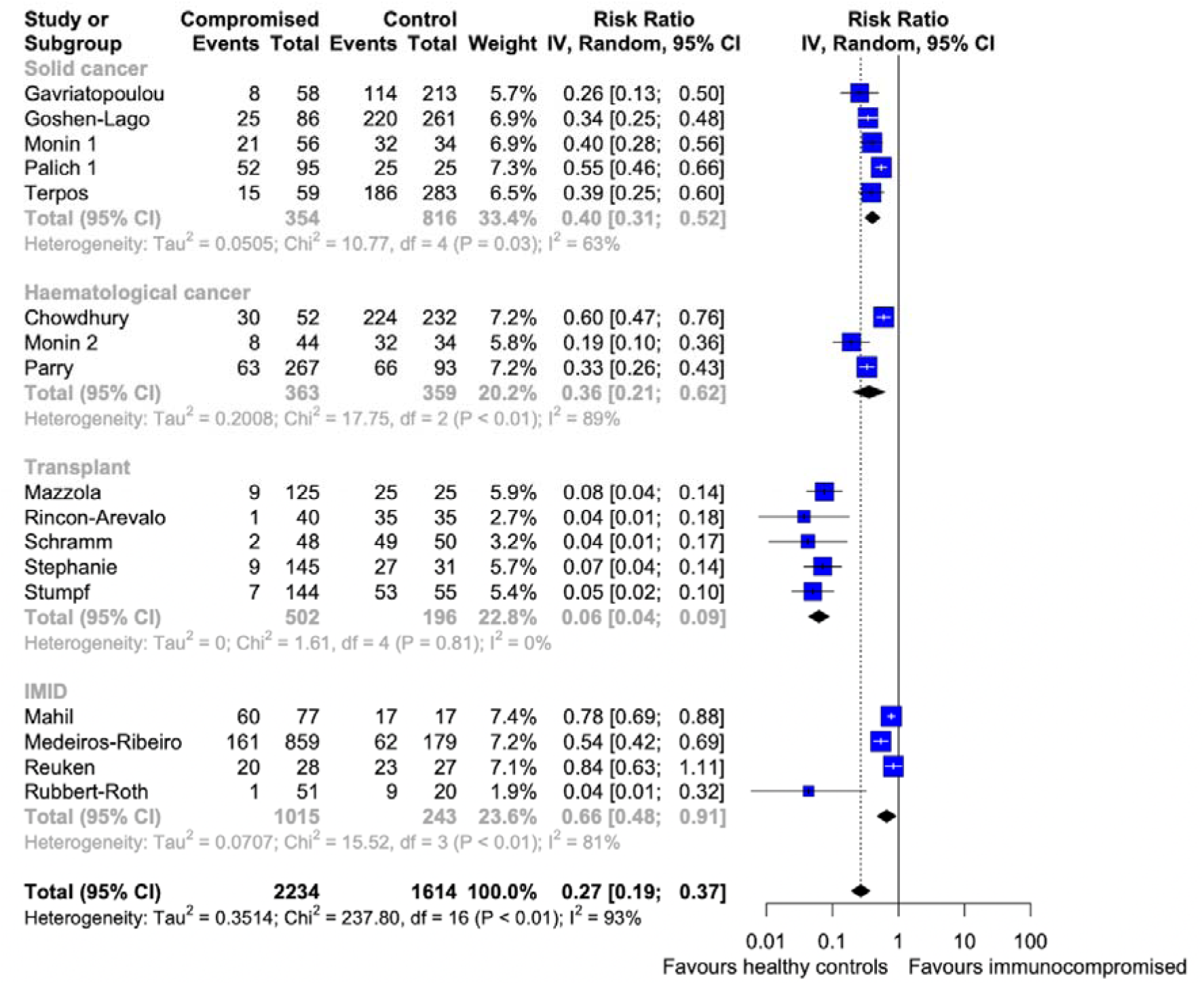
Risk ratio of seroconversion amongst immunocompromised patients compared to healthy controls after first dose

A total of 2234 patients from 18 immunocompromised cohorts in 17 studies were identified reporting seroconversion in immunocompromised patients compared to healthy controls after the first dose. Meta-analysis showed markedly reduced seroconversion rates among the immunocompromised patients as seen by a statistically significant pooled RR of 0.27 (95% confidence interval [CI], 0.19 to 0.37; I^2=93%, p<0.01).

Among the immunocompromised groups, the transplant recipients had the lowest pooled RR with minimal heterogeneity of 0.06 (95%CI: 0.04 to 0.09, I^2=0%, p=0.81) (GRADE = Moderate), followed by patients with haematological cancer at 0.36 (95%CI: 0.21 to 0.62, I^2 = 89%, p<0.01) (GRADE = Moderate) and then patients with solid cancers at 0.40 (95%CI: 0.31 to 0.52, I^2 = 63%, p=0.03) (GRADE = Moderate). The highest pooled RR after the first dose was seen in IMID patients at 0.66 (95%CI: 0.48 to 0.91, I^2=81%, p<0.01) (GRADE = Moderate).

### Dose 2

The proportion with seroconversion and risk ratio of seroconversion amongst immunocompromised patients and healthy controls after the second dose of COVID-19 vaccine are presented in Figure 4 and Figure 5 respectively.

**Figure 4:**
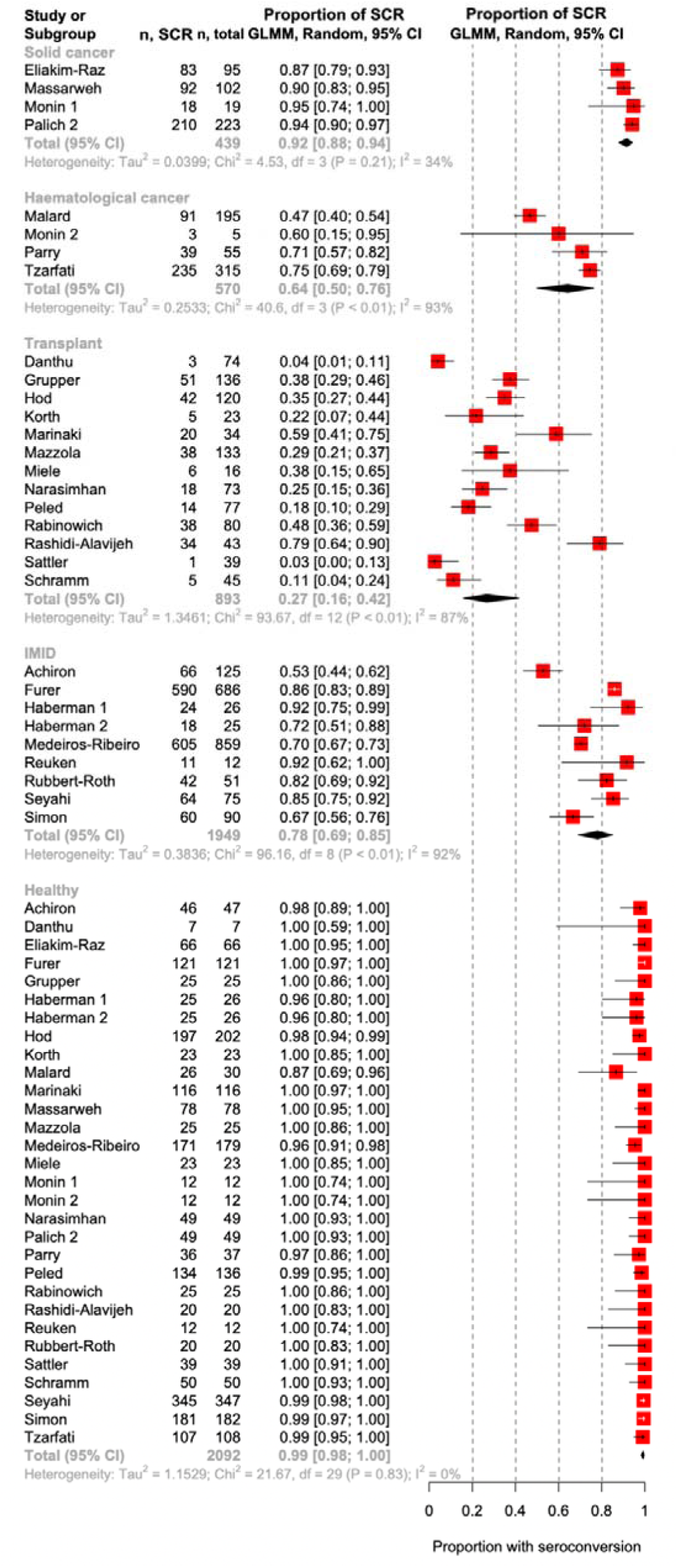
Proportion with seroconversion after the second dose of COVID-19 vaccine; SCR: Seroconversion

**Figure 5:**
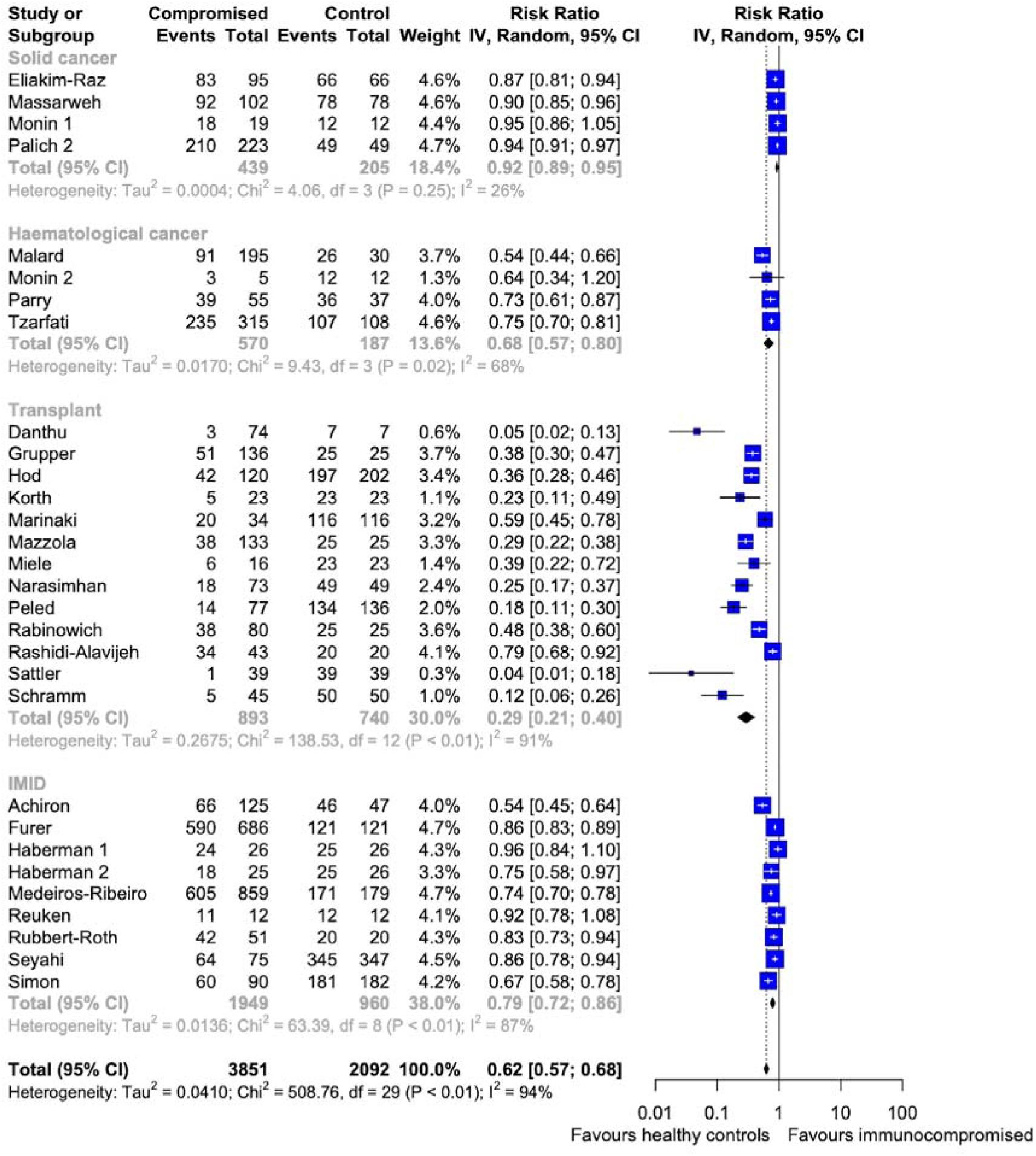
Risk ratio of seroconversion amongst immunocompromised patients compared to healthy controls after second dose

A total of 3851 patients from 30 immunocompromised cohorts in 28 studies were identified reporting seroconversion in immunocompromised patients compared to 2092 healthy controls after the second dose. Meta-analysis of these studies showed a statistically significant pooled RR of 0.62 (95%CI: 0.57 to 0.68, I^2=94%, p<0.01) among the immunocompromised patients. This shows a much higher pooled RR than the immunocompromised patients after the first dose of vaccine, implying the second dose of vaccine greatly boosts the immune response to the vaccine. However, this still shows reduced seroconversion rates in the immunocompromised patients, as compared to healthy controls with a pooled RR of 0.99 (95%CI: 0.98 to 1.00, I^2=0%, p=0.83).

Among the immunocompromised groups, the lowest pooled RR after the second dose was seen in transplant recipients at 0.29 (95%CI: 0.21 to 0.40, I^2=91%, p<0.01) (GRADE = Moderate), followed by haematological cancer at 0.68 (95%CI: 0.57 to 0.80, I^2=68%, p=0.02) (GRADE = Low) and IMID patients at 0.79 (95%CI: 0.72 to 0.86, I^2=87%, p<0.01) (GRADE = Low). The highest pooled RR after the second dose was seen in patients with solid cancer at 0.92 (95%CI: 0.89 to 0.95, I^2=26%, p=0.25) (GRADE = Low).

### Vaccine response in patients with solid cancer after first dose

In cancer patients specifically, interim data has shown lower immune efficacy rates for COVID-19 vaccinations than healthy controls.(63) Immune efficacy of a single inoculum was low in solid cancer patients (<40% efficacious) and even lower in haematological cancer patients (<15%) as compared to healthy controls (>90% efficacious). The impact of immunocompromised states, including malignancies and primary and secondary immunodeficiencies, on the efficacy and immunogenicity of active vaccination is well established in literature on vaccinology. Lower seroconversion and seroprotection was noted among cancer patients receiving influenza vaccination in previous studies.(16)

5 studies including 354 patients with solid cancers and 816 healthy controls showed a pooled RR of 0.40 (95%CI: 0.31 to 0.52).

Terpos et al. 2021(22) provided insights into how immunotherapy could impact antibody titres post COVID vaccination. Antibody titres were 1.73-fold lower (median: 22%; IQR:13.4 to 30.2%) among 59 patients receiving immunotherapy than 283 healthy control subjects (Median: 38%; IQR: 23 to 54%) after the first dose. Similarly, Goshen-Lago et. al 2021(28) showed a 1.70 -fold reduction in antibody titres among 232 patients receiving active treatment for cancer (median: 42.3 AU/mL) compared to 261 healthy controls (median: 72.0 AU/mL).

### Vaccine response in patients with solid cancer after second dose

4 studies including 439 patients with solid cancers and 205 healthy controls showed a pooled RR of 0.92 (95%CI: 0.89 to 0.95) in terms of seroconversion after second dose.

Massarweh et al. 2021(25) demonstrated the most significant reduction of 3.71-fold among 102 patients with (Median: 1931 AU/mL; IQR: 509 to 4386 AU/mL) as compared to 78 healthy control subjects (Median: 7160AU/mL; IQR: 3129 to 11241 AU/mL) after the second dose. The smallest reduction of 2.93-fold was reported by Eliakim-Raz et al.(26), who noted a 2.93-fold reduction among 95 patients with solid cancer (Median: 417 AU/mL; IQR: 136 to 895 AU/mL) as compared to the 66 healthy controls (median: 1220 AU/mL; IQR: 588 to1987 AU/mL).

Seroconversion rates in patients with solid cancer were depressed after both doses, with a significant increase of 2.30 times between the first dose (Pooled RR: 0.40; 95%CI: 0.31 to 0.52) and the second dose (Pooled RR: 0.92; 95%CI: 0.89 to 0.95).

### Vaccine response in patients with haematological cancer after first dose

3 studies including 363 patients with haematological cancers and 359 healthy controls showed a pooled RR of 0.36 (95%CI: 0.21 to 0.62).

Chowdhury et al. 2021(34) reported a significant reduction in antibody titres of 8.4-fold (median: 75 AU/mL; IQR: 19 to 328) among 59 patients with chronic myeloid neoplasms compared to 232 healthy control subjects (median: 630 AU/mL; IQR: 284 to 1328 AU/mL) after the first dose. Parry et al. 2021(33) reported a much higher reduction in antibody titres of 95-fold (median: 0.4U/mL) among 299 patients with chronic lymphocytic leukemia compared to 93 healthy controls (median: 41.6 U/mL). However, Pimpinelli et al.(30) reported antibody titres to be less depressed, with a 2.28-fold (median: 7.5 AU/mL; 95%CI: 5.6 to 10.4 AU/mL) and 1.06-fold (median: 16.2 AU/mL; 95%CI 11.7 to 22.3 AU/mL) reduction amongst multiple myeloma and myeloproliferative neoplasm patients respectively.

### Vaccine response in patients with haematological cancer after second dose

4 studies including 570 patients with haematological cancers and 187 healthy controls showed a pooled RR of 0.68 (95%CI: 0.57 to 0.80) in terms of seroconversion after second dose.

Herishanu et al.(29) demonstrated the most significant reduction of 1315.5-fold among 52 patients with chronic lymphocytic leukemia (median: 0.824U/mL; IQR: 0.4 to 167.3U/mL) as compared to 52 healthy control subjects (median 1084U/mL; IQR: 128.9 to 1879U/mL) after the second dose. The smallest reduction was reported by Tzarfati et al.(31), who noted a 1.85-fold reduction among 315 patients with hematological malignancies (median: 85 AU/mL; IQR: 10.7 to 172AU/mL) as compared to the 108 healthy controls (median: 157 AU/mL; IQR: 130 to 221 AU/mL).

Seroconversion rates in patients with haematological cancer were depressed after both doses, with a significant increase of 1.89 times between the first dose (Pooled RR: 0.36; 95%CI: 0.21 to 0.62) and the second dose (Pooled RR: 0.68; 95%CI: 0.57 to 0.80).

### Vaccine response in patients with rheumatic and other autoimmune conditions after first dose

Rheumatic and other autoimmune disorders often result in potentially life-long immunosuppression, through disease or iatrogenically. Disease modifying anti-rheumatic drugs (DMARDs) such as methotrexate, mycophenolate and biologics targeting B cells are often employed alone or in combination.(13) Being immune-mediated disorders, the immunosuppressive effect of these drugs are used to repress these diseases, though simultaneously compromising vaccine efficacy.(64, 65)

5 studies reported seroconversion rates among IMID patients after the first dose. Data from 1015 IMID patients compared to 243 healthy controls showed that these patients had a pooled RR of 0.66 (95%CI: 0.48 to 0.91).

Lower antibody titres were seen after the first dose of vaccine among IMID patients. Rubbert-Roth et al.(52) demonstrated that the antibody titres of 51 patients with rheumatoid arthritis (median: 0.4 U/mL; IQR: 0.4 to 2.13 U/mL) were much lower than the 20 healthy controls (median: 99.2 U/mL; IQR: 24.8 to 172 U/mL) by 248-fold. Medeiros-Ribeiro et al.(58) showed a less significant decrease in 859 IMID patients (median: 5.1 AU/mL; IQR: 4.7 to 5.5 AU/mL) of 2.02-fold as compared to the 179 healthy controls (median: 10.3 AU/mL; IQR: 8.5 to 12.5 AU/mL).

### Vaccine response in IMID patients after second dose

9 cohorts from 8 studies also reported seroconversion rates among IMID patients after the second dose. Data from 1949 patients with IMID patients compared to 960 healthy controls showed that IMID patients had a pooled RR of 0.79 (95%CI: 0.72 to 0.86).

The greatest and smallest reduction in antibody titres among IMID patients are reported by Achiron et al.(54) Achiron et al. provided insights into the impact of individual immunosuppressive agents on titres of patients with multiple sclerosis 1 month after their second dose. The greatest reduction in antibody titres were seen in the 26 and 44 patients on fingolimod and ocrelizumab respectively, with 27.41 (median: 0.27; IQR: 0.12 to 0.45) and 25.52-fold (median: 0.29; IQR: 0.06 to 0.89) reduction in titres noted respectively compared to 47 healthy control subjects (median: 7.4; IQR: 6.4 to 8.1). The smallest reduction of 0.91-fold in antibody titres was demonstrated among 23 patients on cladribine (median: 7.0; IQR: 6.5 to 8.1) with a 1.06-fold reduction in titres.

Seroconversion rates in IMID patients were depressed after both doses, with a significant increase of 1.20 times between the first dose (Pooled RR: 0.66; 95%CI: 0.48 to 0.91) and the second dose (Pooled RR: 0.79; 95%CI: 0.72 to 0.86).

### Vaccine response in transplant recipients after first dose

5 studies reported seroconversion after the first dose of COVID-19 vaccine in transplant recipients including 502 transplants compared with 196 healthy controls. The depression in seroconversion rates was significant with the pooled RR at 0.06 (95%CI: 0.04 to 0.09) and largely homogenous between studies (I^2=0%, p=0.81).

Mazzola et al.(38) reported a median titre of 153 AU/mL (IQR: 129 to 860) 28 days after the dose of COVID-19 vaccine. However, serological titres of healthy controls were not reported, thus comparison was not possible. Schramm et al.(46) reported only 2 patients with an antibody response 21 days after the first dose. Neither mean nor median serological titres were reported.

### Vaccine response in transplant recipients after second dose

Across 13 studies, pooling 893 transplant recipients and 740 healthy controls, a strong risk for non-seroconversion was found as seen by the lowest pooled RR of 0.29 (95%CI: 0.21 to 0.40).

11 separate studies reported titre levels post-second vaccine dose. Narasimhan et al.(44) reported the greatest fold difference of 8358.24 between transplant recipients and healthy controls, with titres of 1.7 AU/mL (95%CI: 0.6 to 7.5) and 14209 AU/mL (95%CI: 11261 to 18836) respectively. Rabinowich et al.(37) reported the smallest fold reduction in titres of 2.10-fold (Mean±SD: 95.41±92.4 AU/mL vs 200.5±65.1 AU/mL).

Seroconversion rates in transplant recipients were depressed after both doses, with a significant increase of 4.83 times between the first dose (Pooled RR: 0.06; 95%CI: 0.04 to 0.09) and the second dose (Pooled RR: 0.29; 95%CI: 0.21 to 0.40).

From this, it is highly suggestive that a second dose of COVID-19 vaccine is imperative in improving seroconversion rates in transplant recipients. Seroconversion rates remain severely depressed compared to healthy individuals however, thus necessitating future study for third doses or booster shots for such patients. Non-vaccine protective measures would also be vital in protecting this vulnerable group of patients.

### Risk of bias assessment

32 studies were assessed to be at low risk of bias while 11 studies were deemed to be at moderate risk of bias (Supplementary table 2). No studies were at severe or critical risk of bias. Risk of bias mainly came from confounding effects with controls not being age-matched.

### Accounting for heterogeneity in cancer patients after first and second dose

Given the heterogeneity present in the analyses, we undertook subgroup analysis for cancer patients after the first and second dose. We noted that for the first dose, variables like average age, timepoints, brand of serology kit and country of study may account for heterogeneity (Supplementary table 3). However, for the second dose, we did not find any variable that may account for the heterogeneity in data. (Supplementary table 5).

### Publication bias and trim-and-fill analysis for cancer patients after first dose

Egger’s test did not show publication bias in cancer patients (p = 0.9626). Trim-and-fill funnel plot with imputation of potentially missing studies for cancer patients after the first dose (Supplementary figure 1) similarly did not suggest significant bias. 8 studies were combined (with no added studies) with random effects model yielding a result of 0.3142 (95%CI: 0.2201 to 0.4267).

### Publication bias and trim-and-fill analysis for cancer patients after second dose

Similar to the first dose, Egger’s test did not show publication bias in cancer patients (p = 0.2306). Trim-and-fill funnel plot with imputation of potentially missing studies for cancer patients (Supplementary figure 2) combined 11 studies (with 3 added studies) with random effects model yielding a proportion of 0.6813 (95%CI: 0.4709 to 0.8370).

## DISCUSSION

In this systematic review and meta-analysis of 42 studies which included immunocompromised groups of patients with solid cancer, patients with haematological malignancies, transplant recipients and IMID patients, we found that these patients had depressed seroconversion after the first dose and second dose compared to healthy controls. Compared to the pooled RR of 0.91 after the first dose among healthy controls, the pooled RR after the first dose was much lower at 0.06 among transplant recipients, 0.36 among patients with hematological cancers, 0.40 among patients with solid cancers, and 0.66 among IMID patients. Antibody response improved significantly after the second dose. The pooled RR after the second dose increased to 0.29 among transplant recipients, 0.68 among patients with hematological cancers, 0.79 among IMID patients, 0.92 among patients with solid cancers and 0.99 among healthy controls. Transplant recipients demonstrated sustained low seroconversion rates after both doses of vaccine.

To the best of our knowledge, this is the first meta-analysis to study immunogenicity and serologic titre response in immunocompromised patients to the first and second dose of COVID vaccines, stratifying results by the different aetiologies of immunosuppression.

Our findings highlight the importance of the second dose of COVID-19 vaccines and subsequent booster shots. It is well established in literature the benefits of additional doses and boosters of vaccines, both for COVID-19(66–68) and pre-existing vaccines (e.g. inactivated polio vaccine).(69) This review similarly highlights the importance of a second dose vaccine especially for the immunocompromised individuals. Across the included studies, a second dose of vaccine confers greatly improved seroconversion and titre levels. In particular, the administration of a second dose is of great importance in increasing immunogenicity and protection in organ transplant and haematological patients.

### Benefits of a third dose of COVID-19 vaccine

In transplant recipients and patients with haematological malignancies, our results have demonstrated a less-than-ideal seroconversion rate even after a second inoculum, prompting the need for additional measures. Particularly, transplant recipients exhibited a severely depressed seroconversion rate across all studies.

A recent randomised trial studying the immunogenicity of a third dose of the mRNA-1273 (Moderna) vaccine in organ transplant recipients showed a statistically significant benefit of a third dose. 55% of patients in the group receiving a third dose achieved an anti-receptor binding domain antibody level of at least 100U/mL, compared to 18% in those which received a placebo instead.(70) Another study by Del Bello et al.(71) using 3 doses of the BNT162b2 vaccine (Pfizer-BioNTech) vaccine in transplant recipients found that seroconversion rates increased with every dose, from 5.1% (95%CI, 3.0% to 7.4%; n = 20) before the second dose to 41.4% (95%CI, 36.5% to 46.3%; n = 164) before the third dose and finally to 67.9% (95%CI, 63.3% to 72.6%; n = 269) 4 weeks after the third dose. Other studies have had similar studies, proving the effectiveness of a third dose.(72)

The Food and Drug Authority in August of 2021 has authorised the use of a third dose of Pfizer-BioNTech and Moderna vaccines for immunocompromised populations, including transplant recipients(73) with other countries following suit.(74)

Furthermore, our meta-analyses suggest that between aetiologies of immunocompromise, significant heterogeneity in immunogenicity is noted both post-first dose and post-second dose. This suggests that vaccine regimens should be tailored according to the aetiology and degree of immunocompromise. Achiron et al.(54), one of the included studies, also demonstrated significantly different seroconversion rates for those on different therapies. This is supported by Kennedy et al.(75) which underscored the fact that immunosuppression caused by different biologic agents could be substantial; 20 patients on infliximab had significantly lowered titres compared to 7 patients on vedolizumab (mean±SD: 158±7.0U/mL vs 562±11.5U/mL). Ligumsky et al.(76) further demonstrated that different anti-cancer therapies can also lead to varying seroconversion rates and antibody titres with patients on chemotherapy having a lower median IgG titre and seroconversion rate than those on immune checkpoint inhibitors and targeted therapy.

### Surrogate measures of immunogenicity and efficacy

Currently, there is no international consensus on measures to determine immunogenicity. Trials reported surrogate measures including seroconversion rates and geometric mean titres. These surrogate measures involved parameters related to anti-SARS-COV-2 recombinant spike, receptor binding domain or neutralising IgG or total antibodies. The immunological markers and their respective use in predicting protection against COVID-19 has been the subject of much debate.(77–80) While neutralising antibody level has more recently been established to be a reliable predictor of protection against symptomatic COVID-19, it remains that many studies have utilised varying measures.

In this review, only studies which involved comparison of measures of effect to that of immunocompetent controls were included.

### Understudied populations

To date, there is minimal published data comparing the immunogenicity of the COVID-19 vaccine in patients with HIV and AIDS to that in healthy persons. Impairing the T-cell lines, both humoral and cell-based immunity is implicated in HIV and AIDS. Past studies have established lower rates of response to hepatitis B and influenza vaccines in HIV and AIDS.(16, 81–83)

Current published data from a randomised controlled trial in South Africa of patients with HIV demonstrated that immunogenicity was not significantly impaired.(84) Given the HIV-AIDS spectrum, the effect of level of depletion of CD4 counts on immune response remains to be established. A search of the clinical trial register ClinicalTrials.gov yielded multiple upcoming and ongoing studies including HIV and AIDS patients.(85–88)

There is also a paucity of evidence in patients with primary immunodeficiencies.(89) However, even patients with primary antibody deficiencies such as combined variable immunodeficiency have been demonstrated to develop anti-spike antibodies post-vaccination.(90) Hence, all patients with primary immunodeficiencies should be vaccinated against COVID-19.

### Limitations

Firstly, the studies included in this paper are observational studies. Factors that may influence the immune response to the vaccine, such as comorbidities and age, may not be controlled between both the immunocompromised populations and healthy controls. To address this limitation, we performed subgroup analyses which showed no significant effect modification between studies with different median age.

Secondly, there was heterogeneity in the definition of immunocompromised state. To address this limitation, we have pre-specified the definition of immunocompromised and performed subgroup analyses accordingly to assess the difference in seroconversion rates in different groups of immunocompromised patients which revealed stark differences between solid cancer, haematological cancer, IMID and transplant patients.

Next, while seroconversion rates is an indication of the immune response to the vaccine, it is only a proxy for the different impact that the vaccine has on the infection rates and severity of COVID.

Lastly, the definition of seroconversion and immunoassay used are not standardised across the studies. To address this limitation, we have performed subgroup analyses to determine if there is effect modification between studies that used different brands of immunoassays. Interestingly, significant effect modification was shown in dose 1 but not dose 2.

Furthermore, vaccination type may influence the seroconversion rates of individuals after their COVID-19 vaccination. However, given that the studies included in this review predominantly used mRNA vaccines, analyses of possible differences could not be performed.

## CONCLUSION

In this meta-analysis, we have shown that seroconversion rates and serological titres are significantly lower in immunocompromised patients compared to immunocompetent individuals. Among the various groups of immunocompromised patients, organ transplant recipients had the lowest, while solid cancer patients had the highest seroconversion rates. Of note, immunocompromised patients who seroconvert generally develop lower antibody titres than healthy controls, which poses a concern of whether they have indeed achieved an adequate level of seroprotection. Additional strategies on top of the conventional 2-dose regimen for mRNA COVID-19 vaccines would be warranted to confer improved seroprotection for these patients, such as the administration of a third dose.

## SUMMARY

What is already known on this topic

- Immunocompromised patients exhibit lower seroconversion rates than healthy persons after receiving other vaccines, such as the influenza vaccine, but less is known about the response to COVID-19 vaccines, particularly, mRNA-based vaccines.

What this study adds

- This systematic review and network meta-analysis provides a comprehensive overview and evaluation of the evidence published as of 3 September 2021 and will be re-evaluated and updated periodically
- There is moderate certainty that seroconversion in immunocompromised patients after a first dose of COVID-19 vaccine is low.
- While there is a significant increase in seroconversion rates between the first and second dose of COVID-19 vaccine, seroconversion rates still remain depressed among all immunocompromised groups. There is moderate certainty that seroconversion in transplant recipients remain severely depressed even after a second dose.
- Amongst immunocompromised groups studied, antibody titres are lower than in healthy persons.

## CONTRIBUTIONS

ARYBL and SYW contributed equally to this paper and are joint first authors. RS and YYS contributed equally to this paper and are joint corresponding authors. ARYBL, SYW, RS and YYS contributed to study concept and design. ARYBL and SYW selected the articles and extracted the data. BKJT, CBT, YHC, ARYBL and YYS were responsible for statistical analysis. ARYBL and SYW wrote the first draft of the manuscript. RS, YYS, LYAC, SCL, MDM, SHT and ML provided advice at different stages. All authors approved the final version of the manuscript. RS is the guarantor. The corresponding authors attest that all listed authors meet authorship criteria and that no others meeting the criteria have been omitted.

## ETHICAL APPROVAL

Not applicable. All the work was developed using published data.

## DATA SHARING

This study has no additional data available.

## LICENSE FOR PUBLICATION

The Corresponding Author has the right to grant on behalf of all authors and does grant on behalf of all authors, a worldwide licence to the Publishers and its licensees in perpetuity, in all forms, formats and media (whether known now or created in the future), to i) publish, reproduce, distribute, display and store the Contribution, ii) translate the Contribution into other languages, create adaptations, reprints, include within collections and create summaries, extracts and/or, abstracts of the Contribution, iii) create any other derivative work(s) based on the Contribution, iv) to exploit all subsidiary rights in the Contribution, v) the inclusion of electronic links from the Contribution to third party material where-ever it may be located; and, vi) licence any third party to do any or all of the above.

## Data Availability

This study has no additional data available.

## REFERENCES

1. Ali MAM, Spinler SA. COVID-19 and thrombosis: From bench to bedside. Trends Cardiovasc Med. 2021;31(3):143–60.

2. Berlin DA, Gulick RM, Martinez FJ. Severe Covid-19. N Engl J Med. 2020;383(25):2451–60.

3. Fathizadeh H, Afshar S, Masoudi MR, Gholizadeh P, Asgharzadeh M, Ganbarov K, et al. SARS-CoV-2 (Covid-19) vaccines structure, mechanisms and effectiveness: A review. Int J Biol Macromol. 2021;188:740–50.

4. Polack FP, Thomas SJ, Kitchin N, Absalon J, Gurtman A, Lockhart S, et al. Safety and Efficacy of the BNT162b2 mRNA Covid-19 Vaccine. N Engl J Med. 2020;383(27):2603–15.

5. Voysey M, Clemens SAC, Madhi SA, Weckx LY, Folegatti PM, Aley PK, et al. Safety and efficacy of the ChAdOx1 nCoV-19 vaccine (AZD1222) against SARS-CoV-2: an interim analysis of four randomised controlled trials in Brazil, South Africa, and the UK. The Lancet. 2021;397(10269):99–111.

6. Eyre DW, Lumley SF, Wei J, Cox S, James T, Justice A, et al. Quantitative SARS-CoV-2 anti-spike responses to Pfizer-BioNTech and Oxford-AstraZeneca vaccines by previous infection status. Clin Microbiol Infect. 2021.

7. Covid-19 vaccines for moderately to severely immunocompromised people [Internet]. Centers for Disease Control and Prevention. Centers for Disease Control and Prevention; [cited 27 September 2021]. Available from: https://www.cdc.gov/coronavirus/2019-ncov/vaccines/recommendations/immuno.html

8. Manuel O, Estabrook M, American Society of Transplantation Infectious Diseases Community of P. RNA respiratory viral infections in solid organ transplant recipients: Guidelines from the American Society of Transplantation Infectious Diseases Community of Practice. Clin Transplant. 2019;33(9):e13511.

9. Couch MDRB, Englund MDJA. Respiratory Viral Infections in Immunocompetent and Immunocompromised Persons. The American Journal of Medicine. 1997;102(3):2–9.

10. Mohammed AH, Blebil A, Dujaili J, Rasool-Hassan BA. The Risk and Impact of COVID-19 Pandemic on Immunosuppressed Patients: Cancer, HIV, and Solid Organ Transplant Recipients. AIDS Rev. 2020;22(3):151–7.

11. Madan A, Siglin J, Khan A. Comprehensive review of implications of COVID-19 on clinical outcomes of cancer patients and management of solid tumors during the pandemic. Cancer Med. 2020;9(24):9205–18.

12. Pereira MR, Mohan S, Cohen DJ, Husain SA, Dube GK, Ratner LE, et al. COVID-19 in solid organ transplant recipients: Initial report from the US epicenter. Am J Transplant. 2020;20(7):1800–8.

13. Curtis JR, Johnson SR, Anthony DD, Arasaratnam RJ, Baden LR, Bass AR, et al. American College of Rheumatology Guidance for COVID-19 Vaccination in Patients With Rheumatic and Musculoskeletal Diseases: Version 3. Arthritis Rheumatol. 2021.

14. Chong PP, Handler L, Weber DJ. A Systematic Review of Safety and Immunogenicity of Influenza Vaccination Strategies in Solid Organ Transplant Recipients. Clin Infect Dis. 2018;66(11):1802–11.

15. Westra J, van Assen S, Wilting KR, Land J, Horst G, de Haan A, et al. Rituximab impairs immunoglobulin (Ig)M and IgG (subclass) responses after influenza vaccination in rheumatoid arthritis patients. Clin Exp Immunol. 2014;178(1):40–7.

16. Beck CR, McKenzie BC, Hashim AB, Harris RC, University of Nottingham I, the ImmunoCompromised Study G, et al. Influenza vaccination for immunocompromised patients: systematic review and meta-analysis by etiology. J Infect Dis. 2012;206(8):1250–9.

17. Page MJ, McKenzie JE, Bossuyt PM, Boutron I, Hoffmann TC, Mulrow CD, et al. The PRISMA 2020 statement: an updated guideline for reporting systematic reviews. BMJ. 2021;372:n71.

18. Sterne JA, Hernan MA, Reeves BC, Savovic J, Berkman ND, Viswanathan M, et al. ROBINS-I: a tool for assessing risk of bias in non-randomised studies of interventions. BMJ. 2016;355:i4919.

19. Sterne JAC, Savovic J, Page MJ, Elbers RG, Blencowe NS, Boutron I, et al. RoB 2: a revised tool for assessing risk of bias in randomised trials. BMJ. 2019;366:l4898.

20. Guyatt GH, Oxman AD, Vist GE, Kunz R, Falck-Ytter Y, Alonso-Coello P, et al. GRADE: an emerging consensus on rating quality of evidence and strength of recommendations. BMJ. 2008;336(7650):924–6.

21. Monin L, Laing AG, Muñoz-Ruiz M, McKenzie DR, Del Molino Del Barrio I, Alaguthurai T, et al. Safety and immunogenicity of one versus two doses of the COVID-19 vaccine BNT162b2 for patients with cancer: interim analysis of a prospective observational study. Lancet Oncol. 2021;22(6):765–78.

22. Terpos E, Zagouri F, Liontos M, Sklirou AD, Koutsoukos K, Markellos C, et al. Low titers of SARS-CoV-2 neutralizing antibodies after first vaccination dose in cancer patients receiving checkpoint inhibitors. J Hematol Oncol. 2021;14(1):86.

23. Palich R, Veyri M, Marot S, Vozy A, Gligorov J, Maingon P, et al. Weak immunogenicity after a single dose of SARS-CoV-2 mRNA vaccine in treated cancer patients. Ann Oncol. 2021;32(8):1051–3.

24. Palich R, Veyri M, Vozy A, Marot S, Gligorov J, Benderra MA, et al. High seroconversion rate but low antibody titers after two injections of BNT162b2 (Pfizer-BioNTech) vaccine in patients treated with chemotherapy for solid cancers. Ann Oncol. 2021;32(10):1294–5.

25. Massarweh A, Eliakim-Raz N, Stemmer A, Levy-Barda A, Yust-Katz S, Zer A, et al. Evaluation of Seropositivity Following BNT162b2 Messenger RNA Vaccination for SARS-CoV-2 in Patients Undergoing Treatment for Cancer. JAMA Oncol. 2021;7(8):1133–40.

26. Eliakim-Raz N, Massarweh A, Stemmer A, Stemmer SM. Durability of Response to SARS-CoV-2 BNT162b2 Vaccination in Patients on Active Anticancer Treatment. JAMA Oncol. 2021.

27. Gavriatopoulou M, Terpos E, Kastritis E, Briasoulis A, Gumeni S, Ntanasis-Stathopoulos I, et al. Low neutralizing antibody responses in WM, CLL and NHL patients after the first dose of the BNT162b2 and AZD1222 vaccine. Clin Exp Med. 2021.

28. Goshen-Lago T, Waldhorn I, Holland R, Szwarcwort-Cohen M, Reiner-Benaim A, Shachor-Meyouhas Y, et al. Serologic Status and Toxic Effects of the SARS-CoV-2 BNT162b2 Vaccine in Patients Undergoing Treatment for Cancer. JAMA Oncol. 2021.

29. Herishanu Y, Avivi I, Aharon A, Shefer G, Levi S, Bronstein Y, et al. Efficacy of the BNT162b2 mRNA COVID-19 vaccine in patients with chronic lymphocytic leukemia. Blood. 2021;137(23):3165–73.

30. Pimpinelli F, Marchesi F, Piaggio G, Giannarelli D, Papa E, Falcucci P, et al. Fifth-week immunogenicity and safety of anti-SARS-CoV-2 BNT162b2 vaccine in patients with multiple myeloma and myeloproliferative malignancies on active treatment: preliminary data from a single institution. J Hematol Oncol. 2021;14(1):81.

31. Herzog Tzarfati K, Gutwein O, Apel A, Rahimi-Levene N, Sadovnik M, Harel L, et al. BNT162b2 COVID-19 vaccine is significantly less effective in patients with hematologic malignancies. Am J Hematol. 2021;96(10):1195–203.

32. Malard F, Gaugler B, Gozlan J, Bouquet L, Fofana D, Siblany L, et al. Weak immunogenicity of SARS-CoV-2 vaccine in patients with hematologic malignancies. Blood Cancer J. 2021;11(8):142.

33. Parry H, McIlroy G, Bruton R, Ali M, Stephens C, Damery S, et al. Antibody responses after first and second Covid-19 vaccination in patients with chronic lymphocytic leukaemia. Blood Cancer J. 2021;11(7):136.

34. Chowdhury O, Bruguier H, Mallett G, Sousos N, Crozier K, Allman C, et al. Impaired antibody response to COVID-19 vaccination in patients with chronic myeloid neoplasms. Br J Haematol. 2021;194(6):1010–5.

35. Yi SG, Knight RJ, Graviss EA, Moore LW, Nguyen DT, Ghobrial RM, et al. Kidney Transplant Recipients Rarely Show an Early Antibody Response Following the First COVID-19 Vaccine Administration. Transplantation. 2021;105(7):e72–e3.

36. Grupper A, Rabinowich L, Schwartz D, Schwartz IF, Ben-Yehoyada M, Shashar M, et al. Reduced humoral response to mRNA SARS-CoV-2 BNT162b2 vaccine in kidney transplant recipients without prior exposure to the virus. Am J Transplant. 2021;21(8):2719–26.

37. Rabinowich L, Grupper A, Baruch R, Ben-Yehoyada M, Halperin T, Turner D, et al. Low immunogenicity to SARS-CoV-2 vaccination among liver transplant recipients. J Hepatol. 2021;75(2):435–8.

38. Mazzola A, Todesco E, Drouin S, Hazan F, Marot S, Thabut D, et al. Poor Antibody Response after Two Doses of SARS-CoV-2 vaccine in Transplant Recipients. Clin Infect Dis. 2021.

39. Sattler A, Schrezenmeier E, Weber UA, Potekhin A, Bachmann F, Straub-Hohenbleicher H, et al. Impaired humoral and cellular immunity after SARS-CoV-2 BNT162b2 (tozinameran) prime-boost vaccination in kidney transplant recipients. J Clin Invest. 2021;131(14).

40. Marinaki S, Adamopoulos S, Degiannis D, Roussos S, Pavlopoulou ID, Hatzakis A, et al. Immunogenicity of SARS-CoV-2 BNT162b2 vaccine in solid organ transplant recipients. Am J Transplant. 2021;21(8):2913–5.

41. Miele M, Busa R, Russelli G, Sorrentino MC, Di Bella M, Timoneri F, et al. Impaired anti-SARS-CoV-2 humoral and cellular immune response induced by Pfizer-BioNTech BNT162b2 mRNA vaccine in solid organ transplanted patients. Am J Transplant. 2021;21(8):2919–21.

42. Peled Y, Ram E, Lavee J, Sternik L, Segev A, Wieder-Finesod A, et al. BNT162b2 vaccination in heart transplant recipients: Clinical experience and antibody response. J Heart Lung Transplant. 2021;40(8):759–62.

43. Hod T, Ben-David A, Olmer L, Levy I, Ghinea R, Mor E, et al. Humoral Response of Renal Transplant Recipients to the BNT162b2 SARS-CoV-2 mRNA Vaccine Using Both RBD IgG and Neutralizing Antibodies. Transplantation. 2021.

44. Narasimhan M, Mahimainathan L, Clark AE, Usmani A, Cao J, Araj E, et al. Serological Response in Lung Transplant Recipients after Two Doses of SARS-CoV-2 mRNA Vaccines. Vaccines (Basel). 2021;9(7).

45. Rashidi-Alavijeh J, Frey A, Passenberg M, Korth J, Zmudzinski J, Anastasiou OE, et al. Humoral Response to SARS-Cov-2 Vaccination in Liver Transplant Recipients-A Single-Center Experience. Vaccines (Basel). 2021;9(7).

46. Schramm R, Costard-Jäckle A, Rivinius R, Fischer B, Müller B, Boeken U, et al. Poor humoral and T-cell response to two-dose SARS-CoV-2 messenger RNA vaccine BNT162b2 in cardiothoracic transplant recipients. Clin Res Cardiol. 2021;110(8):1142–9.

47. Stumpf J, Siepmann T, Lindner T, Karger C, Schwöbel J, Anders L, et al. Humoral and cellular immunity to SARS-CoV-2 vaccination in renal transplant versus dialysis patients: A prospective, multicenter observational study using mRNA-1273 or BNT162b2 mRNA vaccine. Lancet Reg Health Eur. 2021:100178.

48. Rincon-Arevalo H, Choi M, Stefanski AL, Halleck F, Weber U, Szelinski F, et al. Impaired humoral immunity to SARS-CoV-2 BNT162b2 vaccine in kidney transplant recipients and dialysis patients. Sci Immunol. 2021;6(60).

49. Danthu C, Hantz S, Dahlem A, Duval M, Ba B, Guibbert M, et al. Humoral Response after SARS-CoV-2 mRNA Vaccination in a Cohort of Hemodialysis Patients and Kidney Transplant Recipients. J Am Soc Nephrol. 2021;32(9):2153–8.

50. Korth J, Jahn M, Dorsch O, Anastasiou OE, Sorge-Hädicke B, Eisenberger U, et al. Impaired Humoral Response in Renal Transplant Recipients to SARS-CoV-2 Vaccination with BNT162b2 (Pfizer-BioNTech). Viruses. 2021;13(5).

51. Furer V, Eviatar T, Zisman D, Peleg H, Paran D, Levartovsky D, et al. Immunogenicity and safety of the BNT162b2 mRNA COVID-19 vaccine in adult patients with autoimmune inflammatory rheumatic diseases and in the general population: a multicentre study. Ann Rheum Dis. 2021;80(10):1330–8.

52. Rubbert-Roth A, Vuilleumier N, Ludewig B, Schmiedeberg K, Haller C, von Kempis J. Anti-SARS-CoV-2 mRNA vaccine in patients with rheumatoid arthritis. Lancet Rheumatol. 2021;3(7):e470–e2.

53. Deepak P, Kim W, Paley MA, Yang M, Carvidi AB, El-Qunni AA, et al. Glucocorticoids and B Cell Depleting Agents Substantially Impair Immunogenicity of mRNA Vaccines to SARS-CoV-2. medRxiv. 2021.

54. Achiron A, Mandel M, Dreyer-Alster S, Harari G, Magalashvili D, Sonis P, et al. Humoral immune response to COVID-19 mRNA vaccine in patients with multiple sclerosis treated with high-efficacy disease-modifying therapies. Ther Adv Neurol Disord. 2021;14:17562864211012835.

55. Geisen UM, Berner DK, Tran F, Sümbül M, Vullriede L, Ciripoi M, et al. Immunogenicity and safety of anti-SARS-CoV-2 mRNA vaccines in patients with chronic inflammatory conditions and immunosuppressive therapy in a monocentric cohort. Ann Rheum Dis. 2021;80(10):1306–11.

56. Izmirly PM, Kim MY, Samanovic M, Fernandez-Ruiz R, Ohana S, Deonaraine KK, et al. Evaluation of Immune Response and Disease Status in SLE Patients Following SARS-CoV-2 Vaccination. Arthritis Rheumatol. 2021.

57. Mahil SK, Bechman K, Raharja A, Domingo-Vila C, Baudry D, Brown MA, et al. The effect of methotrexate and targeted immunosuppression on humoral and cellular immune responses to the COVID-19 vaccine BNT162b2: a cohort study. Lancet Rheumatol. 2021;3(9):e627–e37.

58. Medeiros-Ribeiro AC, Aikawa NE, Saad CGS, Yuki EFN, Pedrosa T, Fusco SRG, et al. Immunogenicity and safety of the CoronaVac inactivated vaccine in patients with autoimmune rheumatic diseases: a phase 4 trial. Nat Med. 2021.

59. Reuken PA, Andreas N, Grunert PC, Glöckner S, Kamradt T, Stallmach A. T cell response after SARS-CoV-2 vaccination in immunocompromised patients with inflammatory bowel disease. J Crohns Colitis. 2021.

60. Seyahi E, Bakhdiyarli G, Oztas M, Kuskucu MA, Tok Y, Sut N, et al. Antibody response to inactivated COVID-19 vaccine (CoronaVac) in immune-mediated diseases: a controlled study among hospital workers and elderly. Rheumatol Int. 2021;41(8):1429–40.

61. Haberman RH, Herati RS, Simon D, Samanovic M, Blank RB, Tuen M, et al. Methotrexate Hampers Immunogenicity to BNT162b2 mRNA COVID-19 Vaccine in Immune-Mediated Inflammatory Disease. medRxiv. 2021.

62. Simon D, Tascilar K, Fagni F, Kronke G, Kleyer A, Meder C, et al. SARS-CoV-2 vaccination responses in untreated, conventionally treated and anticytokine-treated patients with immune-mediated inflammatory diseases. Ann Rheum Dis. 2021;80(10):1312–6.

63. Monin-Aldama L, Laing AG, Muñoz-Ruiz M, McKenzie DR, del Molino del Barrio I, Alaguthurai T, et al. Interim results of the safety and immune-efficacy of 1 versus 2 doses of COVID-19 vaccine BNT162b2 for cancer patients in the context of the UK vaccine priority guidelines. 2021.

64. Mehta P, Sanchez E, Moraitis E, Longley N, Lendrem DW, Giles IP, et al. Influenza vaccination and interruption of methotrexate in adult patients in the COVID-19 era: an ongoing dilemma. The Lancet Rheumatology. 2021;3(1):e9–e10.

65. Nakafero G, Grainge MJ, Myles PR, Mallen CD, Zhang W, Doherty M, et al. Effectiveness of inactivated influenza vaccine in autoimmune rheumatic diseases treated with disease-modifying anti-rheumatic drugs. Rheumatology (Oxford). 2020;59(12):3666–75.

66. Doria-Rose N, Suthar MS, Makowski M, O’Connell S, McDermott AB, Flach B, et al. Antibody Persistence through 6 Months after the Second Dose of mRNA-1273 Vaccine for Covid-19. N Engl J Med. 2021;384(23):2259–61.

67. Livingston EH. Necessity of 2 Doses of the Pfizer and Moderna COVID-19 Vaccines. JAMA. 2021;325(9):898.

68. Silva-Cayetano A, Foster WS, Innocentin S, Belij-Rammerstorfer S, Spencer AJ, Burton OT, et al. A booster dose enhances immunogenicity of the COVID-19 vaccine candidate ChAdOx1 nCoV-19 in aged mice. Med (N Y). 2021;2(3):243–62 e8.

69. Pregliasco F, Grilli G, Andreassi A, D’Addezio E, Vacca F, Squarcione S, et al. Immune response to a booster dose of enhanced potency inactivated polio vaccine administered in association with HBV vaccine in adolescents. Vaccine. 1996;14(4):267–9.

70. Hall VG, Ferreira VH, Ku T, Ierullo M, Majchrzak-Kita B, Chaparro C, et al. Randomized Trial of a Third Dose of mRNA-1273 Vaccine in Transplant Recipients. N Engl J Med. 2021;385(13):1244–6.

71. Del Bello A, Abravanel F, Marion O, Couat C, Esposito L, Lavayssiere L, et al. Efficiency of a boost with a third dose of anti-SARS-CoV-2 messenger RNA-based vaccines in solid organ transplant recipients. Am J Transplant. 2021.

72. Benotmane I, Gautier G, Perrin P, Olagne J, Cognard N, Fafi-Kremer S, et al. Antibody Response After a Third Dose of the mRNA-1273 SARS-CoV-2 Vaccine in Kidney Transplant Recipients With Minimal Serologic Response to 2 Doses. JAMA. 2021.

73. Coronavirus (COVID-19) Update: FDA Authorizes Additional Vaccine Dose for Certain Immunocompromised Individuals [Internet]. U.S. Food and Drug Administration. 2021 [cited 27 September 2021]. Available from: https://www.fda.gov/news-events/press-announcements/coronavirus-covid-19-update-fda-authorizes-additional-vaccine-dose-certain-immunocompromised.

74. Wise J. Covid-19: UK will offer third vaccine dose to severely immunosuppressed people. BMJ. 2021;374:n2160.

75. Kennedy NA, Lin S, Goodhand JR, Chanchlani N, Hamilton B, Bewshea C, et al. Infliximab is associated with attenuated immunogenicity to BNT162b2 and ChAdOx1 nCoV-19 SARS-CoV-2 vaccines in patients with IBD. Gut. 2021;70(10):1884–93.

76. Ligumsky H, Safadi E, Etan T, Vaknin N, Waller M, Croll A, et al. Immunogenicity and Safety of the BNT162b2 mRNA COVID-19 Vaccine Among Actively Treated Cancer Patients. J Natl Cancer Inst. 2021.

77. Jin P, Li J, Pan H, Wu Y, Zhu F. Immunological surrogate endpoints of COVID-2019 vaccines: the evidence we have versus the evidence we need. Signal Transduct Target Ther. 2021;6(1):48.

78. Khoury DS, Cromer D, Reynaldi A, Schlub TE, Wheatley AK, Juno JA, et al. Neutralizing antibody levels are highly predictive of immune protection from symptomatic SARS-CoV-2 infection. Nat Med. 2021;27(7):1205–11.

79. Earle KA, Ambrosino DM, Fiore-Gartland A, Goldblatt D, Gilbert PB, Siber GR, et al. Evidence for antibody as a protective correlate for COVID-19 vaccines. Vaccine. 2021;39(32):4423–8.

80. Garcia-Beltran WF, Lam EC, Astudillo MG, Yang D, Miller TE, Feldman J, et al. COVID-19-neutralizing antibodies predict disease severity and survival. Cell. 2021;184(2):476–88 e11.

81. Anema A, Mills E, Montaner J, Brownstein JS, Cooper C. Efficacy of influenza vaccination in HIV-positive patients: a systematic review and meta-analysis. HIV Med. 2008;9(1):57–61.

82. Catherine FX, Piroth L. Hepatitis B virus vaccination in HIV-infected people: A review. Hum Vaccin Immunother. 2017;13(6):1–10.

83. Lee JH, Hong S, Im JH, Lee JS, Baek JH, Kwon HY. Systematic review and meta-analysis of immune response of double dose of hepatitis B vaccination in HIV-infected patients. Vaccine. 2020;38(24):3995–4000.

84. Madhi SA, Koen AL, Izu A, Fairlie L, Cutland CL, Baillie V, et al. Safety and immunogenicity of the ChAdOx1 nCoV-19 (AZD1222) vaccine against SARS-CoV-2 in people living with and without HIV in South Africa: an interim analysis of a randomised, double-blind, placebo-controlled, phase 1B/2A trial. The Lancet HIV. 2021;8(9):e568–e80.

85. Immunogenicity of COVID-19 Vaccination in PLWH - ClinicalTrials.gov [Internet]. Clinicaltrials.gov. 2021 [cited 27 September 2021]. Available from: https://clinicaltrials.gov/ct2/show/NCT04894448.

86. Antibodies Production After Covid-19 Vaccination Among Patients With Medical History of Cancer and Anti-CD-20 Treatment - ClinicalTrials.gov [Internet]. Clinicaltrials.gov. 2021 [cited 27 September 2021]. Available from: https://clinicaltrials.gov/ct2/show/NCT04779996.

87. Study of the Humoral Response to SARS-CoV-2 Variants and of the Cellular Response After Vaccination Against COVID-19 in Immunocompromised People - ClinicalTrials.gov [Internet]. Clinicaltrials.gov. 2021 [cited 27 September 2021]. Available from: https://clinicaltrials.gov/ct2/show/NCT04844489.

88. COVID-19 CoronaVac in Patients With Autoimmune Rheumatic Diseases and HIV/AIDS - ClinicalTrials.gov [Internet]. Clinicaltrials.gov. 2021 [cited 27 September 2021]. Available from: https://clinicaltrials.gov/ct2/show/NCT04754698.

89. Roifman CM, Vong L. COVID-19 vaccination for patients with primary immunodeficiency. LymphoSign Journal. 2021;8(2):37–45.

90. Hagin D, Freund T, Navon M, Halperin T, Adir D, Marom R, et al. Immunogenicity of Pfizer-BioNTech COVID-19 vaccine in patients with inborn errors of immunity. J Allergy Clin Immunol. 2021;148(3):739–49.

