## Supplementary tables and figures for "Efficacy of COVID-19 vaccines in immunocompromised patients: A systematic review and meta-analysis"

**Supplementary table 2: Risk of bias of all included controlled observational studies using the ROBINS-I scale**

|  | Domain 1: Risk of bias due to confounding | Domain 2: Bias in selection of participants into the study | Domain 3: Bias in classification of interventions | Domain 4: Bias due to deviations from intended interventions | Domain 5: Bias due to missing data | Domain 6: Bias in measurement of outcomes | Domain 7: Bias in selection of the reported result | Overall bias |
| --- | --- | --- | --- | --- | --- | --- | --- | --- |
| Seyahi et al. | L | L | L | L | L | L | L | L |
| Peled et al. | L | L | L | L | L | L | L | L |
| Herishanu et al. | L | L | L | L | L | L | L | L |
| Massarweh et al. | L | L | L | L | L | L | L | L |
| Pimpinelli et al. | L | L | L | L | L | L | L | L |
| Deepak et al. | M | L | L | L | L | L | L | M |
| Achiron et al. | L | L | L | L | L | L | L | L |
| Danthu et al. | L | L | L | L | L | L | L | L |
| Geisen et al. | L | L | L | L | L | L | L | L |
| Furer et al. | L | L | L | L | L | L | L | L |
| Sattler et al. | L | L | L | L | L | L | L | L |
| Rincon-Arevalo et al. | L | L | L | L | L | L | L | L |
| Korth et al. | L | L | L | L | L | L | L | L |
| Haberman et al. | L | L | L | L | L | L | L | L |
| Grupper et al. | L | L | L | L | L | L | L | L |
| Monin et al. | M | L | L | L | L | L | L | M |
| Simon et al. | L | L | L | L | L | L | L | L |
| Rubbert-Roth et al. | M | L | L | L | L | L | L | M |
| Chowdhury et al. | L | L | L | L | L | L | L | L |
| Stephanie et al. | M | L | L | L | L | L | L | M |
| Rabinowich et al. | L | L | L | L | L | L | L | L |
| Terpos et al. | M | L | L | L | L | L | L | M |
| Mazzola et al. | L | L | L | L | L | L | L | L |
| Palich et al. (Dose 1) | L | L | L | L | L | L | L | L |
| Palich et al. (Dose 2) | L | L | L | L | L | L | L | L |
| Marinaki et al. | L | L | L | L | L | L | L | L |
| Miele et al. | M | L | L | L | L | L | L | M |
| Eliakim-Raz et al. | L | L | L | L | L | L | L | L |
| Gavriatopoulou et al. | L | L | L | L | L | L | L | L |
| Goshen-Lago et al. | L | L | L | L | L | L | L | L |
| Tzarfati et al. | L | L | L | L | L | L | L | L |
| Hod et al. | M | L | L | L | L | L | L | M |
| Izmirly et al. | L | L | L | L | L | L | L | L |
| Mahil et al. | L | L | L | L | L | L | L | L |
| Malard et al. | M | L | L | L | L | L | L | M |
| Medeiros-Ribeiro et al. | L | L | L | L | L | L | L | L |
| Narasimhan et al. | M | L | L | L | L | L | L | M |
| Parry et al. | L | L | L | L | L | L | L | L |
| Rashidi-Alavijeh et al. | M | L | L | L | L | L | L | M |
| Reuken et al. | L | L | L | L | L | L | L | L |
| Schramm et al. | L | L | L | L | L | L | L | L |
| Stumpf et al. | M | L | L | L | L | L | L | M |

L: Low risk of bias; M: Moderate risk of bias

**Supplementary table 3: Mixed effects meta-regression of log(HRs) against potential effect moderators (continuous and categorical study-level characteristics) for the longitudinal association of cancer status with seroconversion proportion after first dose of COVID-19 vaccine**

|  | **Beta‡** | **SE** | **Z** | **P** | **95% CI Lower** | **95% CI Upper** | **I^2^ (% residual heterogeneity)** |
| --- | --- | --- | --- | --- | --- | --- | --- |
| Average age | -0.1310 | 0.0441 | -2.9685 | 0.0030* | -0.2176 | -0.0445 | 73.6894% |
| Vaccine type | 0.4487 | 0.6978 | 0.6430 | 0.5202 | -0.9190 | 1.8163 | 83.4540% |
| ROBINS-I of moderate | -0.1357 | 0.5886 | -0.2306 | 0.8176 | -1.2893 | 1.0179 | 88.2460% |
| Timepoint of 2-4 weeks | -0.9562 | 0.4690 | -2.0387 | 0.0415* | -1.8754 | -0.0369 | 82.0388% |
| Brand of serology kit | | | | | | | |
| GenScript | -1.6401 | 0.2858 | -5.7382 | <0.0001* | -2.2003 | -1.0799 | 0.0000% |
| Roche | -1.4073 | 0.2199 | -6.3999 | <0.0001* | -1.8383 | -0.9763 |  |
| Countries | | | | | | | |
| Greece | -1.6386 | 0.6092 | -2.6899 | 0.0071* | -2.8326 | -0.4447 | 71.5583% |
| Israel | -1.0927 | 0.6798 | -1.6074 | 0.1080 | -2.4250 | 0.2396 |  |
| UK | -0.9140 | 0.5355 | -1.7067 | 0.0879 | -1.9636 | 0.1356 |  |

Abbreviations: HR, hazard ratio; ROBINS-I, Risk Of Bias In Non-randomized Studies of Interventions; CI, confidence interval

‡ Estimated factor by which the log(HR) changes per unit increase in a continuous variable or in comparison with the reference group for a categorical variable. 95% CIs are also presented in log scale.

**Supplementary table 4: Meta-analyses of seroconversion incidence and risk ratio of seroconversion compared to immunocompetent individuals after the first dose of COVID-19 vaccine in subgroups, stratified by categorical study-level characteristics**

|  | **Seroconversion incidence** | | | | **Risk ratio of seroconversion** | | |
| --- | --- | --- | --- | --- | --- | --- | --- |
|  | **Studies** | **Incidence (95% CI)** | | **I^2^** | **Studies** | **HR (95% CI)** | **I^2^** |
| **Overall** | 17 | 0.20 (0.11-0.33) | | 94% | 17 | 0.04 (0.02-0.08) | 89% |
| **Population** | | | | | | | |
| Cancer | 8 | 0.31 (0.22-0.42) | | 88% | 8 | 0.08 (0.05-0.14) | 57% |
| Organ transplant | 5 | 0.06 (0.04-0.08) | | 0% | 5 | 0.00 (0.00-0.01) | 33% |
| IMID | 4 | 0.32 (0.06-0.79) | | 98% | 4 | 0.24 (0.07-0.69) | 60% |
| Healthy | 16 | 0.91 (0.79-0.96) | | 93% | Reference | | |
| **Subgroup analysis of cancer patients** | | | | | | | |
| **Variable** | **Studies** | | **Incidence (95% CI)** | | | **I^2^** | |
| **Brand of serology kit (Cancer)** | | | | | | | |
| GenScript | 2 | | 0.20 (0.14-0.29) | | | 59% | |
| Roche | 1 | | 0.24 (0.19-0.29) | | | - | |
| Abbott | 2 | | 0.56 (0.48-0.64) | | | 0% | |
| **Vaccine type (Cancer)** | | | | | | | |
| mRNA | 7 | | 0.32 (0.21-0.46) | | | 87% | |
| Inactivated | 1 | | 0.24 (0.19-0.29) | | | - | |
| **Country (Cancer)** | | | | | | | |
| UK | 4 | | 0.33 (0.20-0.49) | | | 89% | |
| Greece | 2 | | 0.20 (0.14-0.29) | | | 59% | |
| France | 1 | | 0.55 (0.45-0.64) | | | - | |
| Israel | 1 | | 0.29 (0.20-0.39) | | | - | |

**Supplementary figure 1: Funnel plot with trim-and-fill imputation of potentially missing studies after first dose in cancer patients**

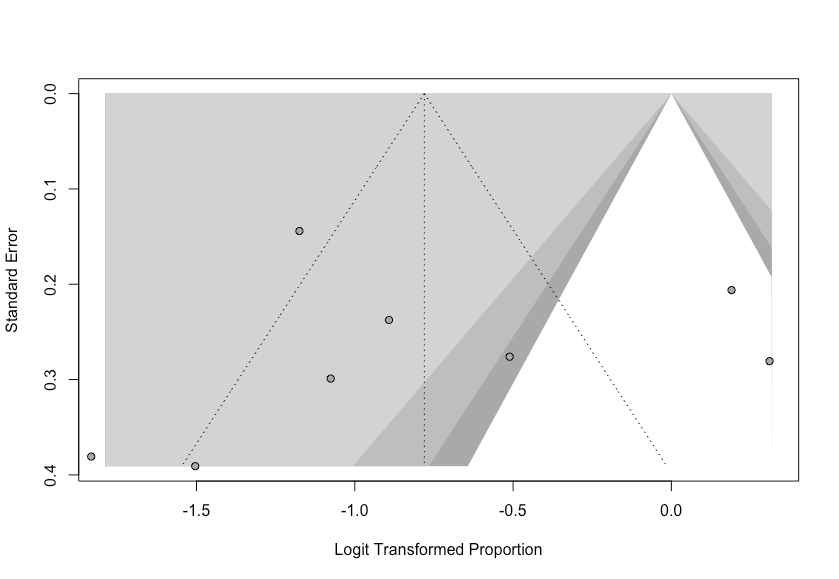

**Trim-and-fill sensitivity analysis**

Number of studies combined: k = 8 (with 0 added studies)

Using random effects model

Proportion: 0.3142

95%-CI: 0.2201; 0.4267

Quantifying heterogeneity:

tau^2 = 0.4106; tau = 0.6408; I^2 = 87.9% [78.4%; 93.2%]; H = 2.88 [2.15; 3.84]

Test of heterogeneity:

Q: 57.86

d.f.: 7

P-value < 0.0001

**Egger’s test for publication bias**

p-value = 0.9626

**Supplementary table 5: Mixed effects meta-regression of log(HRs) against potential effect moderators (continuous and categorical study-level characteristics) for the longitudinal association of cancer status with seroconversion proportion after second dose of COVID-19 vaccine**

|  | **Beta‡** | **SE** | **Z** | **P** | **95% CI Lower** | **95% CI Upper** | **I^2^ (% residual heterogeneity)** |
| --- | --- | --- | --- | --- | --- | --- | --- |
| Average age | -0.1207 | 0.1312 | -0.9204 | 0.3574 | -0.3778 | 0.1364 | 91.6355% |
| Vaccine type | 0.6993 | 1.0508 | 0.6655 | 0.5057 | -1.3602 | 2.7589 | 93.3685% |
| ***ROBINS-I of moderate*** | -0.9697 | 0.6974 | -1.3905 | 0.1644 | -2.3365 | 0.3972 | 87.8956% |
| ***Timepoint of 2-4 weeks*** | 1.0549 | 0.6849 | 1.5403 | 0.1235 | -0.2874 | 2.3973 | 0.0000% |
| Brand of serology kit | | | | | | | |
| LIAISON | -0.2220 | 0.9495 | -0.2338 | 0.8151 | -2.0830 | 1.6389 | 90.8000% |
| Roche | -0.3962 | 0.9873 | -0.4014 | 0.6882 | -2.3312 | 1.5387 |  |
| Country | | | | | | | |
| Israel | 0.4530 | 0.8765 | 0.5169 | 0.6052 | -1.2648 | 2.1709 | 90.5017% |
| UK | 0.0955 | 0.9494 | 0.1006 | 0.9199 | -1.7653 | 1.9563 |  |

Abbreviations: HR, hazard ratio; ROBINS-I, Risk Of Bias In Non-randomized Studies of Interventions; CI, confidence interval

‡ Estimated factor by which the log(HR) changes per unit increase in a continuous variable or in comparison with the reference group for a categorical variable. 95% CIs are also presented in log scale.

**Supplementary table 6: Meta-analyses of seroconversion incidence and risk ratio of seroconversion compared to immunocompetent individuals after the second dose of COVID-19 vaccine in subgroups, stratified by categorical study-level characteristics**

|  | **Seroconversion incidence** | | | | **Risk ratio of seroconversion** | | |
| --- | --- | --- | --- | --- | --- | --- | --- |
|  | **Studies** | **Incidence (95% CI)** | | **I^2^** | **Studies** | **HR (95% CI)** | **I^2^** |
| **Overall** | 29 | 0.60 (0.46-0.73) | | 96% | 29 | 0.03 (0.02-0.05) | 54% |
| **Population** | | | | | | | |
| Cancer | 8 | 0.82 (0.69-0.90) | | 95% | 8 | 0.09 (0.04-0.19) | 0% |
| Organ transplant | 12 | 0.27 (0.15-0.43) | | 88% | 12 | 0.01 (0.00-0.02) | 0% |
| IMID | 9 | 0.78 (0.69-0.85) | | 92% | 9 | 0.07 (0.03-0.14) | 24% |
| Healthy | 27 | 0.99 (0.98-1.00) | | 0% | Reference | | |
| **Subgroup analysis of cancer patients** | | | | | | | |
| **Variable** | **Studies** | | **Incidence (95% CI)** | | | **I^2^** | |
| **Brand of serology kit** | | | | | | | |
| Roche | 1 | | 0.71 (0.58-0.81) | | | - | |
| Abbott | 3 | | 0.79 (0.52-0.93) | | | 97% | |
| LIAISON | 1 | | 0.75 (0.70-0.79) | | | - | |
| **Vaccine type** | | | | | | | |
| mRNA | 7 | | 0.83 (0.69-0.92) | | | 95% | |
| Inactivated | 1 | | 0.71 (0.58-0.81) | | | - | |
| **Country** | | | | | | | |
| UK | 3 | | 0.79 (0.58-0.91) | | | 49% | |
| France | 2 | | 0.79 (0.33-0.97) | | | 99% | |
| Israel | 3 | | 0.84 (0.75-0.91) | | | 86% | |

Abbreviations: HR, hazard ratio; CI, confidence interval.

HR, 95% CI, and I^2^ for subgroups with only 1 constituent study are not reported and instead indicated with a dash (-). Prediction intervals are reported for subgroups with at least 3 studies.

**Supplementary figure 2: Funnel plot with trim-and-fill imputation of potentially missing studies after second dose in cancer patients**

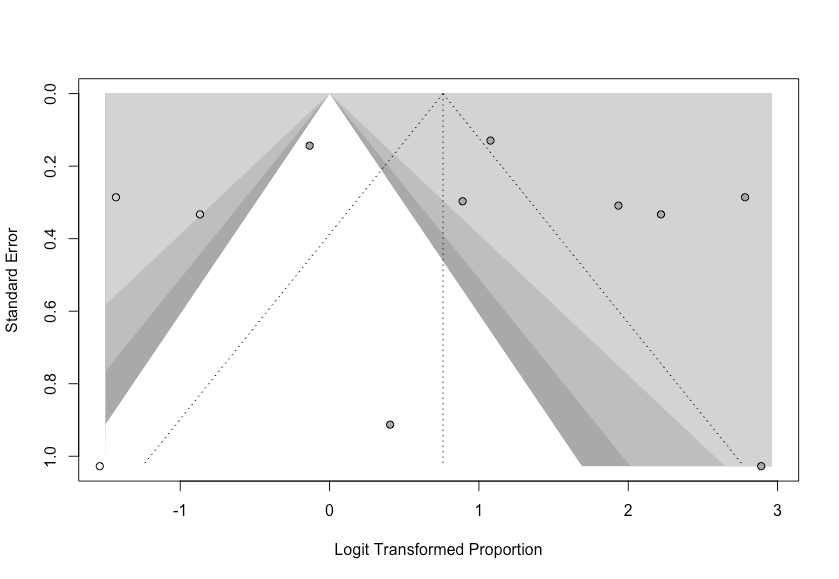

**Trim-and-fill sensitivity analysis**

Number of studies combined: k = 11 (with 3 added studies)

Using random effects model:

Proportion: 0.6813

95%-CI: 0.4709; 0.8370

Quantifying heterogeneity:

tau^2 = 1.9374; tau = 1.3919; I^2 = 95.4% [93.4%; 96.8%]; H = 4.69 [3.90; 5.62]

Test of heterogeneity:

Q: 219.52

d.f.: 10

P-value < 0.0001

**Egger’s test for publication bias**

p-value = 0.2306
